# Optimal Clinical Trials Platform for Progressive Multiple Sclerosis (OCTOPUS): protocol for an international, multi-arm, multi-stage, platform, randomized controlled, double-blind, phase 3 clinical trial

**DOI:** 10.64898/2026.06.15.26355245

**Authors:** Sean Apap Mangion, Charles Wade, Cheryl Pugh, Matthew Burnell, Rachel Burton, Mary Rauchenberger, Hannah Sweeney, Aoife Nolan, Monica Lewis, Elizabeth Brodnicki, Fleur Hudson, Rachael Hunter, Ekaterina Bordea, Rasha Abdel-Fahim, Tarunya Arun, Simon A Broadley, Floriana De Angelis, Anisha Doshi, Peter Foley, Helen Ford, Ian Galea, Joe Guadagno, Charles Hillier, Seema Kalra, Simon Kerrigan, Oliver Leach, Dawn Lyle, Fiona Magill, Miriam Mattoscio, Gavin McDonell, Owen Pearson, Stefano Pluchino, Claire Rice, Basil Sharrack, Eli Silber, Cord Spilker, Bindu Yoga, Amanda Adler, Susan Pavitt, Denise Fitzgerald, Anna Williams, Susan Scott, Sam Loveless, Rod Middleton, Marie Braisher, Olga Ciccarelli, Frederik Barkhof, Siddharthan Chandran, Alan Thompson, Emma Gray, James Carpenter, Emma Tallantyre, Jennifer Nicholas, Mahesh Parmar, Jeremy Chataway

## Abstract

**Introduction:** Current treatments for multiple sclerosis (MS) do not address the pathological processes of neurodegeneration and chronic demyelination. This, coupled with the significant challenges of translating promising phase 2 results to phase 3 trial success, highlights the need for more efficient trial designs, such as platform multi-arm multi-stage (MAMS) trial approaches. MAMS trials have demonstrated success in areas such as oncology and infectious diseases. They are typified by a statistically robust core trial design that allows the addition of further treatment arms and utilisation of interim outcome analyses at pre-defined timepoints, to determine whether to terminate a treatment arm early or proceed to the final outcome analysis.

To address the challenges in progressive multiple sclerosis (PMS) treatment discovery, the Optimal Clinical Trials Platform for PMS (OCTOPUS) trial was developed. It currently utilises MRI whole-brain atrophy as its interim outcome measure and the clinically relevant composite Expanded Disability Status Scale Plus (EDSS-Plus) as its final outcome measure. A rigorous and systematic drug selection process that assessed preclinical in vitro and animal model evidence, along with additional human data, led to the prioritisation of R/S-alpha lipoic acid (R/S-ALA) and metformin for testing against placebo, targeting pathobiological mechanisms relevant to PMS. All participants will be eligible to receive the current standard of care, including disease-modifying treatments (DMTs).

**Method and analysis:** OCTOPUS will be a multi-centre, randomised, placebo-controlled, double-blind, phase 3, MAMS trial of participants aged 25 to 70 years (inclusive) with PMS and an EDSS score of 4.0 to 8.0 (inclusive). Steady progression must be the major cause of increasing disability rather than relapse in the preceding 2 years. In the trial’s first candidate drug cycle, participants will be allocated to R/S-ALA, metformin, or placebo in a 1:1:1 ratio.

Cycle 1 active treatments will start as R/S-ALA 600 mg once daily, increased after 4 weeks to 600 mg twice daily, or metformin 1 g once daily, increased after 4 weeks to 1 g twice daily. The trial will be multinational, with participation from 28 hospitals across the UK and 10 hospitals in Australia.

Clinician-reported measures will include: the EDSS-Plus and the individual components: EDSS, Timed 25 Foot Walk (T25FW); 9 Hole Peg Test (9HPT); Symbol Digit Modalities Test (SDMT); Sloan Low Contrast Visual Acuity (SLCVA); and Relapse assessment. Patient-reported outcomes include MS specific walking, fatigue, pain, and impact scales. We will include a health economic analysis.

Analysis stage 1 will require randomisation of 125 participants per arm and utilise MRI percentage brain volume change (PBVC) with the Structural Image Evaluation using Normalisation of Atrophy (SIENA) technique from baseline to 78 weeks. A positive outcome in analysis stage 1 will detect a 0.15% per year whole brain atrophy difference with a one-sided alpha of 0.35 and power of 95%, ensuring a low probability of erroneously rejecting a treatment arm at this stage. Any arms that show a positive effect will proceed to final analysis stage 2. Analysis stage 2 will require 600 participants per arm. Participants included in stage 1 will also be included in the stage 2. Analysis stage 2 will evaluate time to 6-month confirmed disability progression in the EDSS-Plus, in order to detect a 25% hazard ratio reduction with 90% power and an alpha of 0.05. Assuming one treatment arm proceeds to analysis stage 2, the trial will recruit approximately 1,200 participants and last about 6 years. This is approximately two-thirds the size and half the duration of separately conducted two-arm phase 2 and 3 trials.

**Ethics and dissemination:** The protocol was approved by the London Hampstead REC (22/LO/0622). This manuscript is based on protocol **version 8.0**, 28^th^ August 2025. The findings of this trial will be disseminated through peer-reviewed publications and conference presentations. There will be a close communication strategy developed with the UK MS Society (MSS) and full patient and public involvement and engagement (PPIE).

**Trial registration:** ISRCTN: 14048364

EudraCT number: 2021-003034-37

CTA 20363/0445

IRAS number: 1003943

Secondary identifying numbers: ND001, CPMS 54274

**Strengths and limitations:**

- The OCTOPUS trial will be the first platform multi-arm multi-stage phase 3 trial in PMS, offering the potential to significantly expedite clinical trial processes with advantages in cost- and time-efficiency, focusing specifically on the poorly treated pathobiological processes of chronic neurodegeneration and demyelination
- It will begin by assessing two promising drug candidates, immediate-release metformin and R/S-ALA, and will expand over the duration of the trial to include more drug arms under the same trial master protocol
- The flexible and statistically robust trial design means that several components of the design (such as the early analysis stage 1 interim outcome) can be updated in line with evolving scientific knowledge
- It will ultimately be the largest ever investigator-initiated phase 3 trial in PMS
- It will include a range of national and international trial sites, including neuroscience centres and district general hospitals
- It will have a high inclusion limit for age (up to 70 years) and disability (up to EDSS 8.0)
- Several components (the telephone EDSS and virtual patient-reported outcome measures) will be amenable to remote collection increasing inclusivity and thus addressing public and participant suggestions, while minimising the risk of missing data
- The main challenges in this trial design are the statistical and methodological complexity involved in design and implementation, and interpretation of interim trial results.

**Conclusion:** The trial launched cycle 1 in January 2023. Analysis stage 1 recruitment of 375 participants was achieved in November 2024, enabling planned interim analysis stage 1 to be conducted by late 2026 (*Figure 1*). On the 1^st^ of June 2026, in the UK, 24 sites are active with a further 4 in set-up as part of stage 2, and in the Australian extension, Platform Adaptive Trial for Remyelination and Neuroprotection in Multiple Sclerosis (PLATYPUS), 1 site is active, with 9 additional sites in set-up.

## INTRODUCTION

Multiple sclerosis (MS) is a chronic, inflammatory, demyelinating, neurodegenerative disorder of the central nervous system (CNS)^1^ affecting approximately 150,000 people in the UK^2^, 35,000 in Australia^3^ and around 3 million globally^4^. There are significant associated financial costs for patients, their caregivers, and the National Health Service (NHS), estimated to be between £22,000 and £34,000 per year in the UK for those with greater disability (Expanded Disability Status Scale (EDSS) ≥4.0)^5,6^.The annual cost of MS in Australia is estimated to be over A$3 billion. Despite success in tackling inflammatory activity^7–9^ with over 20 compounds licensed for relapsing remitting MS (RRMS)^10^, neurogeneration and chronic demyelination driven MS disease progression^11,12^ remain without effective treatments and are the core driver of worsening disability in progressive MS (PMS)^12,13^. This highlights the need for faster effective treatment evaluations by more efficient designs across trial phases, with particular potential for platform multi-arm multi-stage (MAMS) trial designs in PMS ^14–16^.

Platform MAMS trials are typified by a core trial design with statistically robust flexible elements, and the ability to add further arms at pre-defined time points as new within-trial cycles (see **Figure 1**). They have proven advantages in terms of time, cost, and participant numbers^16^, and have resulted in changes in clinical practice in various fields including infectious diseases^17,18^ and oncology^19,20^. MAMS trials are now being utilised for neurodegenerative diseases such as motor neuron disease (MND-SMART^21,22^), and Parkinson’s disease (EJS ACT-PD^23^), while the related multi-arm trial design has already been successfully utilised in PMS (the phase 2b 4-arm neuroprotection MS-SMART trial^24^).

**Figure 1.**
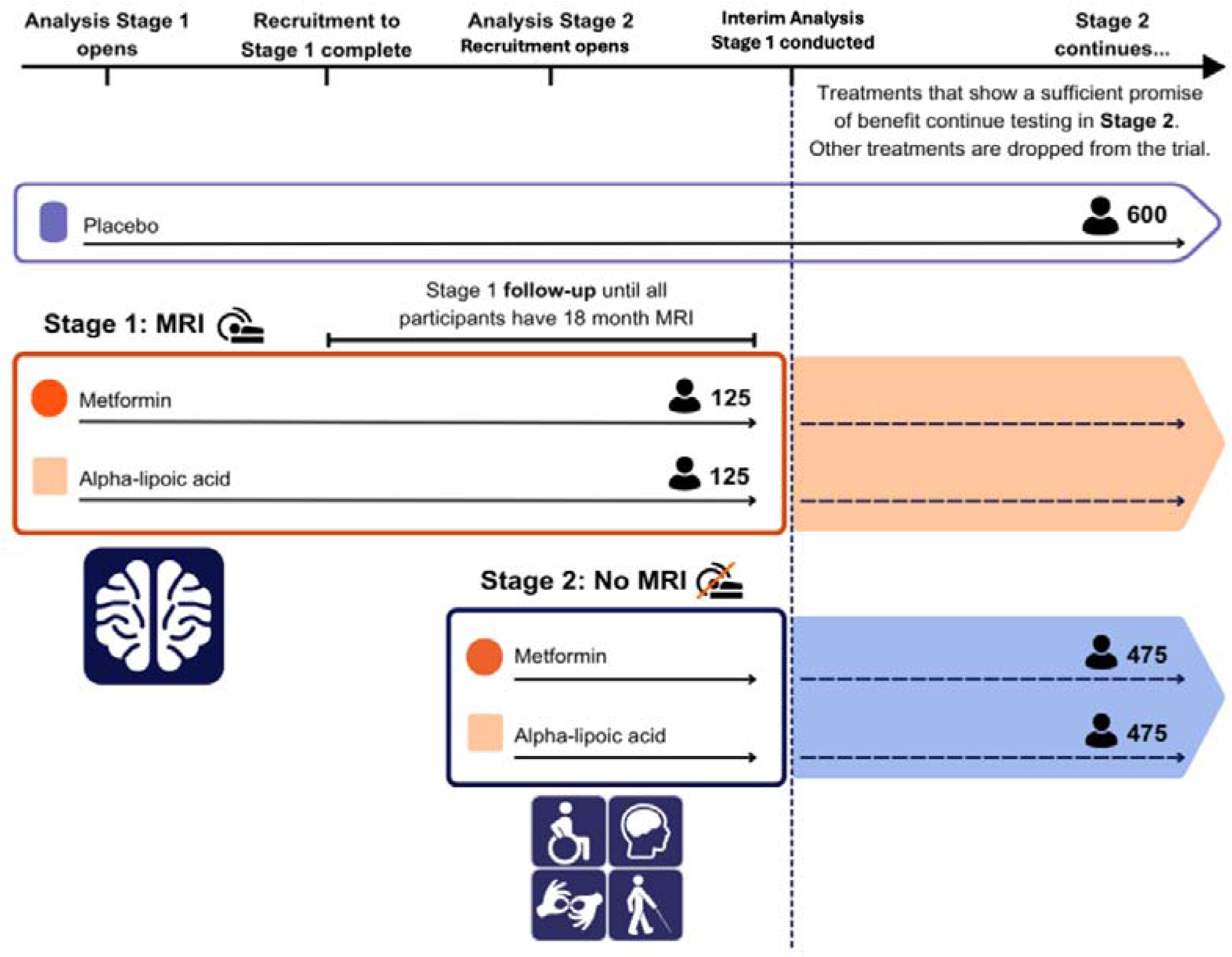
Overview of OCTOPUS MAMS Trial Design. Participant timeline from seamless recruitment across stages 1 and 2, including those that will and will not undertake MRI whole brain atrophy measurements, and interim analysis leading to either arm cessation or continuing recruitment for stage 2. MRI; magnetic resonance imaging

To capitalise on the benefits of MAMS trials, a ready pipeline of potential new treatments is required. For OCTOPUS, a rigorous drug selection process involved the development of an in-depth drug curriculum vitae for shortlisted agents, reflecting facets such as plausibility of biological mechanism of action, quality of early trial/translational evidence, patient preference, and other practical elements^25^. Two key repurposed agents were recommended for further investigation: R- and S-enantiomer racemic mix R/S-alpha lipoic acid (R/S-ALA) and metformin, targeting different direct and indirect mechanisms.

R/S-ALA has in vitro evidence of efficacy across several models, demonstrating nerve cell protection^26–29^, neuronal tissue antioxidant activity^30^ and immune-pathway benefit^31–34^, with further evidence of effect on MS human immune cells^35,36^. Murine in vivo studies also showed benefit for various histological markers as well as for clinical disease onset^31,32,37^, and disease progression^31^. At the time of selection, there was also phase 2 human trial data showing a significant reduction in whole-brain MRI atrophy rates^38^ and benefits for walking performance in people with secondary progressive multiple sclerosis^39^. Metformin has evidence of in vitro preservation of neuronal viability under duress^40,41^, and in vivo murine studies showing benefit on stress-activating cascades^42–44^, oligodendroglial precursor cell numbers and differentiation^44,45^, and immune-mediated pathways^43,44,46^.

## METHODS AND ANALYSES

### Trial Objective

The OCTOPUS design will test drug efficacy compared to placebo by using a two-stage design: analysis stage 1 will use an earlier timepoint **interim** analysis of efficacy, while analysis stage 2 will be the **final** primary outcome analysis. This design will allow an early decision to stop a treatment arm for lack of benefit after stage 1, resulting in considerable time and resource savings and requiring fewer participants ^15,47^. Importantly, the analysis stage 1 outcome measure can be updated to reflect changes in scientific understanding, such as emerging external trial data, but will reflect the underlying processes, clinical change, and treatment effect expected of the analysis stage 2 primary outcome measure. Crucially, analysis stage 1, compared with stage 2, will require a smaller number of participants to be followed up over a shorter duration, with a statistical focus on minimising the type 2 error rate^15^.

Analysis stage 1 will focus on the rate of MRI whole-brain atrophy as the trial’s interim outcome, an established predictor of long-term disease progression in all MS subtypes^48,49^, measured as percentage brain volume change (PBVC) using the SIENA (Structural Image Evaluation, using Normalisation, of Atrophy) technique^50^. The participant numbers and minimum duration of follow-up needed were selected following a review of phase 2 and 3 clinical trials data in PMS, which had also confirmed that treatment effects on clinical outcomes were accompanied by treatment effects on brain atrophy^15^. Atrophy rates up to 104 weeks of follow-up will be compared between treatment and placebo groups in 125 participants per arm, with a one-sided alpha of 0.35 serving as the threshold for a lack of benefit. Treatment arms that demonstrate an effect will continue to analysis stage 2 to be assessed on the final primary outcome.

Analysis stage 2 will assess time to 6-month confirmed disability progression (CDP) in EDSS-Plus as the trial’s final outcome, which is a composite outcome comprising the Expanded Disability Status Scale (EDSS), Timed 25-Foot Walk (T25FW) test, and 9-Hole Peg Test (9HPT). Analysis stage 2 will require 600 participants per arm and aim to detect a hazard ratio of 0.75 for comparison of treatment to to placebo. Assuming one treatment progresses to analysis stage 2, the trial will recruit approximately 1,200 participants (excluding participants from any terminated arm from the same cycle) and last about 6 years. This is approximately two-thirds the size and half the duration of two separate two-arm phase 2 and phase 3 trials ^15^.

### Trial Design and Setting

OCTOPUS will be a randomised, multi-centre, international, double-blinded, multi-arm multi-stage trial, initially comprising three arms, including a single placebo arm for pairwise comparison with two active arms. The initial interventions administered will be either a total daily dose of 50:50 R- and S-enantiomer racemic mix alpha lipoic acid (R/S-ALA) 1200 mg or immediate release metformin 2000 mg, versus placebo, for either pwSPMS or primary progressive MS (PPMS). All participants will be eligible to receive MS treatments as part of standard of care, including DMTs and fampridine.

The participant timeline is illustrated in **Figure 2**, and **Supplementary Table 1** shows the assessment schedule, including the clinician and patient-reported outcomes. Analysis stage 1 of the trial will involve 14 UK National Health Service (NHS) hospitals, including a mix of neuroscience centres and district general hospitals, with plans to expand to more sites once MRI-dependent analysis stage 1 recruitment is concluded. Participating hospitals will be geographically spread between England, Scotland, Wales, and Northern Ireland. Analysis stage 2 will involve analysis stage 1 UK trial sites, as well as further sites spread across the UK and Australia.

**Figure 2.**
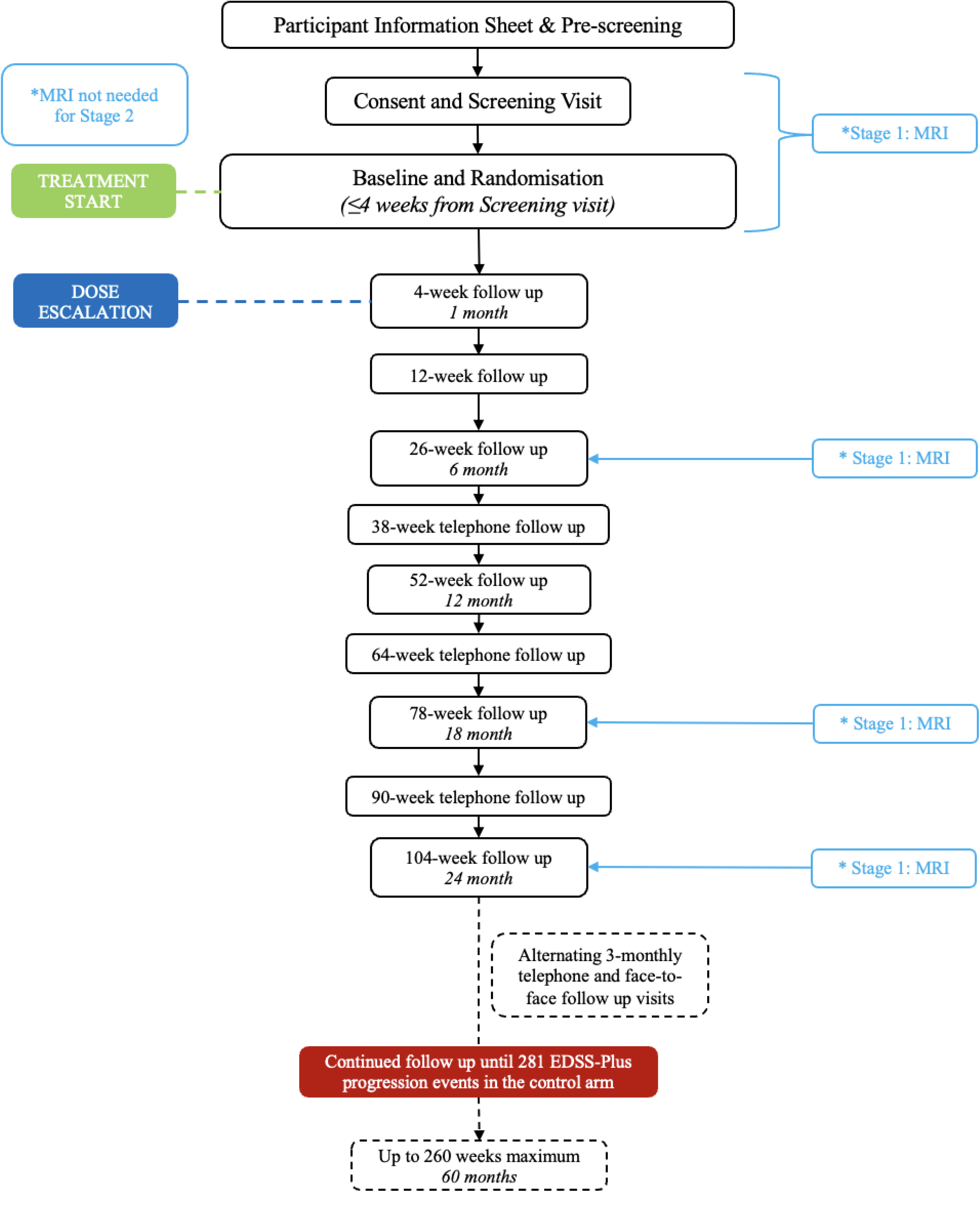
Trial Participant Timeline. Flowsheet of participant timeline from initial contact to conclusion. EDSS-Plus, Expanded Disability Status Scale Plus; MRI, magnetic resonance imaging

### Eligibility criteria

Participants will be aged between 25 and 70 years (inclusive), have a confirmed diagnosis of MS utilising the 2017 McDonald criteria for MS^1^ and have either SPMS or PPMS^1,51^. Increasing disability must be primarily driven by steady progression over the preceding two years, rather than by relapses. The core inclusion and exclusion criteria are described in **Tables 2** and **3**, respectively. In addition to the core criteria, there will be further arm-specific eligibility criteria. Therefore, it will be possible for an individual to be eligible for the trial but not for the currently available arms, in which case they cannot proceed to randomisation in the current trial iteration.

**Table 2.**
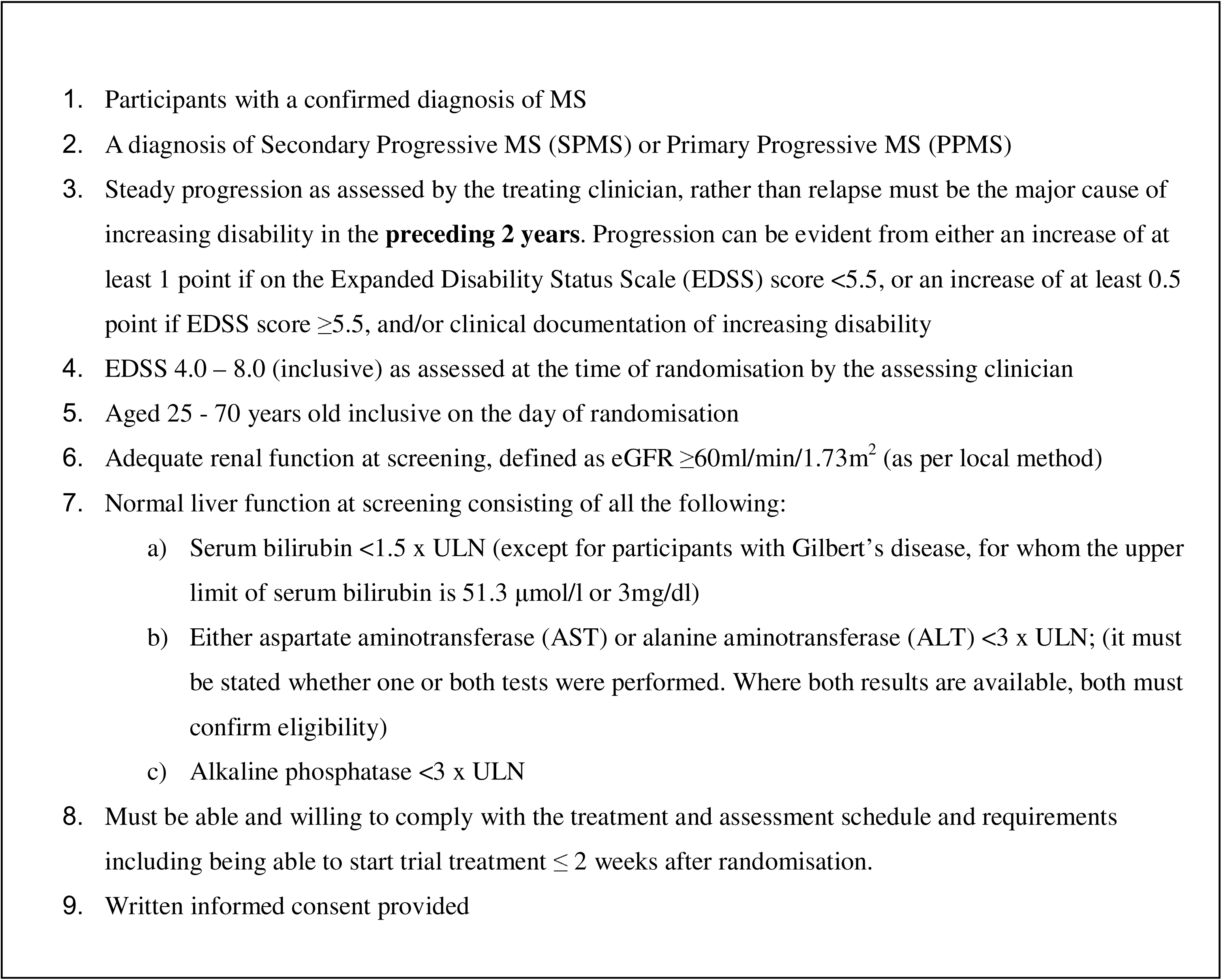
Core Inclusion Criteria.

**Table 3.**
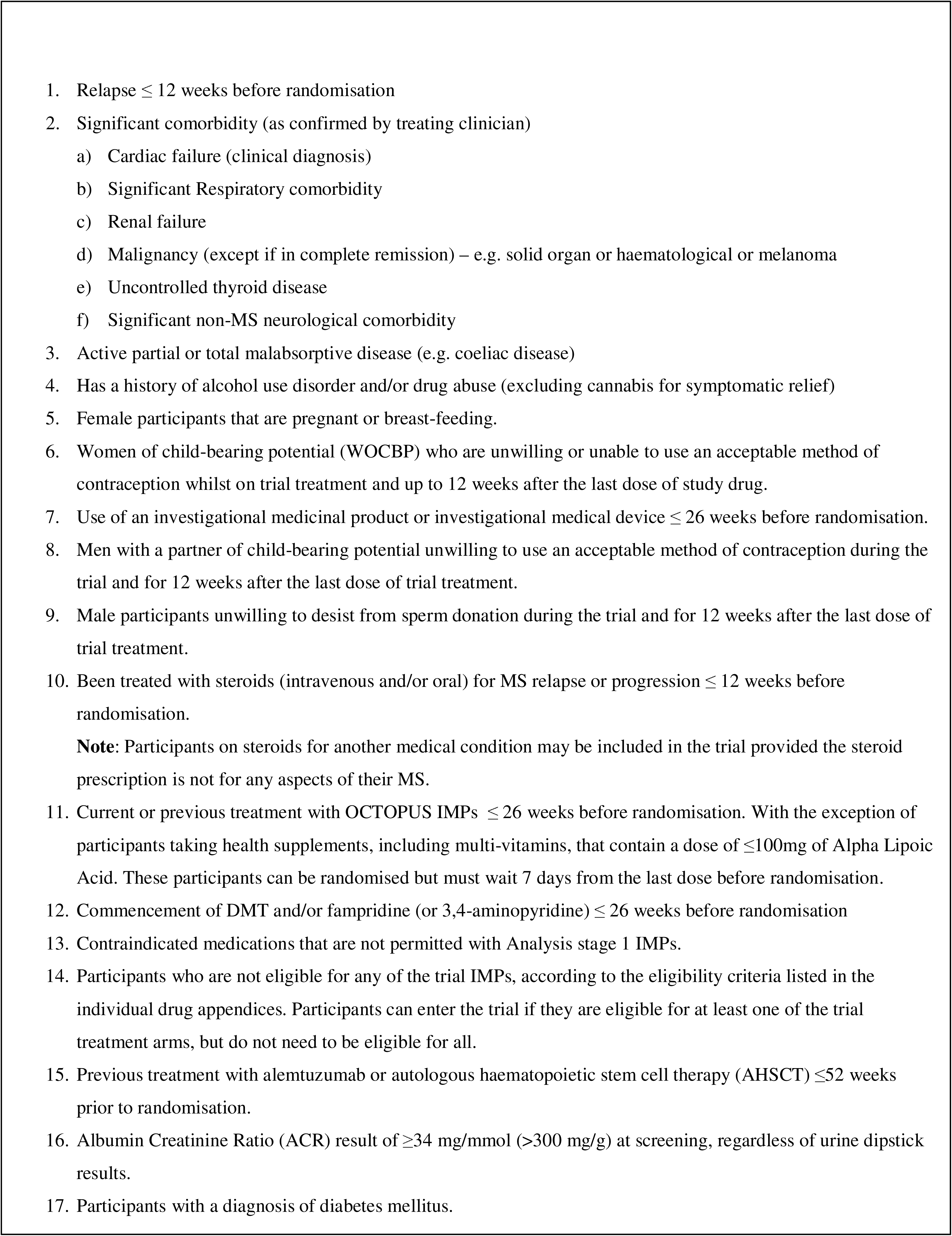
Core Exclusion Criteria.

### Recruitment and Consent

Potential participants will be required to register their interest in joining the trial via a national-level Registration of Interest (ROI) portal (https://ukmsregister.org/octopus). This portal will be freely accessible to all interested participants. After registration, participants will first be invited to share their details (e.g first and last names) and contact details to facilitate further communication with participating sites. They will subsequently be invited to answer a brief set of questions confirming their site preference, diagnosis, and degree of disability. They will also be made aware of trial commitments.

The ROI portal will inform potential participants if they are ineligible from more straightforward questions (for example, an applicant who confirms they have relapsing-remitting MS will be informed that they are not eligible). Participants who are potentially eligible will have their details for more in-depth screening via a phone call with their relevant site.

Participating sites will have experience in MS trials and care, long-term local MS patient population links, and records of patients who may be eligible to participate. During telephone pre-screening of potential participants, sites will check if potential recruites appear to meet core inclusion eligibility. If the potential recruit meets eligibility criteria, and expresses interest to participate, they will be provided with the trial’s Participant Information Sheet (PIS) and a face-to-face trial screening visit scheduled. If potential participants are deemed temporarily ineligible (for example, having recently experienced a relapse, or started a DMT), their details will be stored for later follow-up. At the screening visit, prior to any trial-specific procedures being carried out, participants will be informed of the aims, methods, benefits and potential risks of involvement in the trial. Their understanding of the trial will be confirmed, and they will then be required to give documented informed consent.

### Treatment allocation

In cycle 1, participants will be randomised in a 1:1:1 ratio between R/S-ALA, immediate release metformin, or placebo. Randomisation will be based on a minimisation algorithm with a random element, incorporating: sex (male/female), age (<45 and ≥45), EDSS baseline score 4.0 to 5.5, 6.0 to 6.5, and ≥7.0), use of DMT at baseline (yes/no), PMS phenotype (PPMS/SPMS), and recruiting trial site. Delegated site staff members will verify participant eligibility and consent, then use an independent and secure online randomisation service to directly allocate participants to a treatment arm in a blinded fashion using the participant’s unique identification number. When accessing the drug supply management system to order trial drug or placebo, descriptive information (day, month, and year of birth, and the participant’s unique numeric trial identifier) will be entered to ensure randomisation allocation matches the intended participant.

### Blinding

Site investigators (including pharmacy), the trial sponsor team, and participants will be blinded to treatment allocation. Solely the trial statisticians and randomisation service will have access to unblinded treatment allocations during trial recruitment and follow-up.

Upon randomisation, site staff will be issued with a seven-character code that identifies a blinded bottle of either an active trial drug or placebo according to the participant’s allocation. A hard copy of the code and the prescription will be taken to the site pharmacy for direct issuance to the participant. The same online drug supply management system will be used to order and provide subsequent bottles (‘maintenance kits’) of active trial drug or placebo to the participant on the same basis as at randomisation. It will not be possible to ascertain from kit codes which treatment arm a participant, or group of participants, at any site is on.

All participants will be unblinded once the statistical data lock and final analysis of their arm has occurred. This could be at analysis stage 1 if a treatment is not found to be effective and the arm is terminated, or analysis stage 2. The treatment allocations of all participants randomised by site will be conveyed to the respective site’s Principal Investigator in writing. It will be the responsibility of the Principal Investigator or delegate to inform participants of their treatment allocation, where considered appropriate. There will be a procedure in place for emergency unblinding (detailed in **Monitoring; *Emergency contacts and unblinding***).

## INTERVENTION

### Form

Oral R/S-ALA 300 mg, immediate-release metformin hydrochloride 500 mg, or placebo, over-encapsulated capsules. The R/S-ALA will be manufactured by Pure Encapsulations LLC (Massachusetts, United States of America), and metformin will be manufactured by Relonchem (Cheshire, United Kingdom). Both IMPs will be purchased, over-encapsulated, packaged, labelled, and distributed by Sharp Clinical Services (Wales, United Kingdom). In order to maintain blinding, the over-encapsulating capsules will be identical in appearance for placebo and both IMP preparations; microcrystalline cellulose will fill the capsule for placebo, and back-fill the R/S-ALA and metformin preparations, such that placebo, R/S-ALA, and metformin be identical in appearance and weight.

### Dosage

Participants will receive either R/S-ALA 600 mg twice daily (initially 600 mg once daily at randomisation, then escalated after 4 weeks), metformin 1000 mg twice daily (initially 1000 mg once daily at randomisation, then escalated after 4 weeks), or the same quantity of placebo (of the same colour/size), taken as two capsules twice daily (once daily for the first 4 weeks).

### Duration

Participants will continue on their allocated treatment arm for up to 260 weeks. Participants who discontinue treatment will continue to be followed up unless they withdraw consent to participate in the trial.

### Modification and stopping

The dose may be varied or stopped for patients experiencing defined arm-specific expected adverse events (for current active trial drug expected adverse events see **Table 4**), or other adverse events at the Principal Investigator’s discretion. Re-challenge to a higher dose or restarting treatment will be permissible twice for the same adverse event within up to 4 weeks of reduction from the high dose, if it occurs within 26 weeks post-randomisation. After 26 weeks, the medication dose can only be reduced or stopped.

**Table 4.**
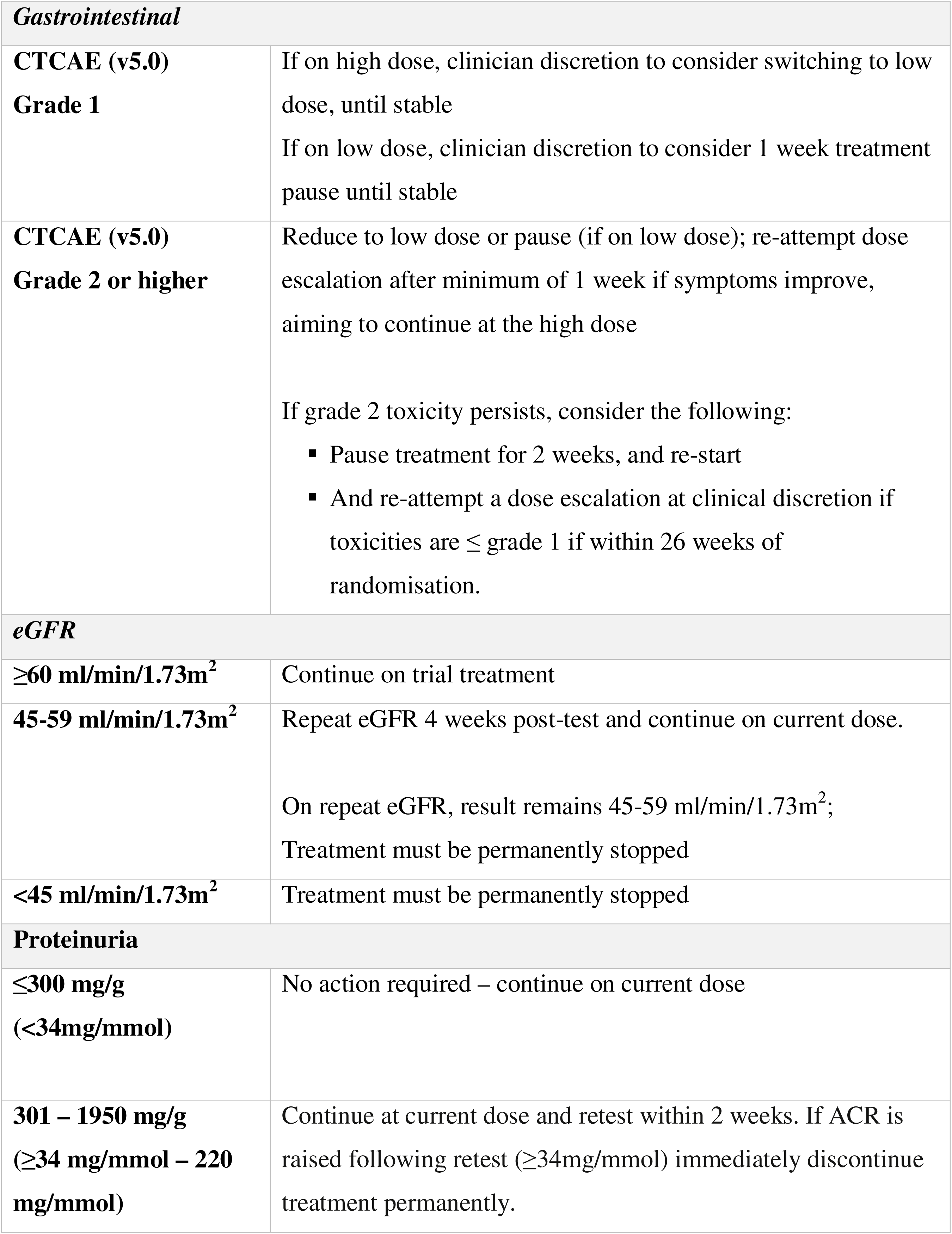
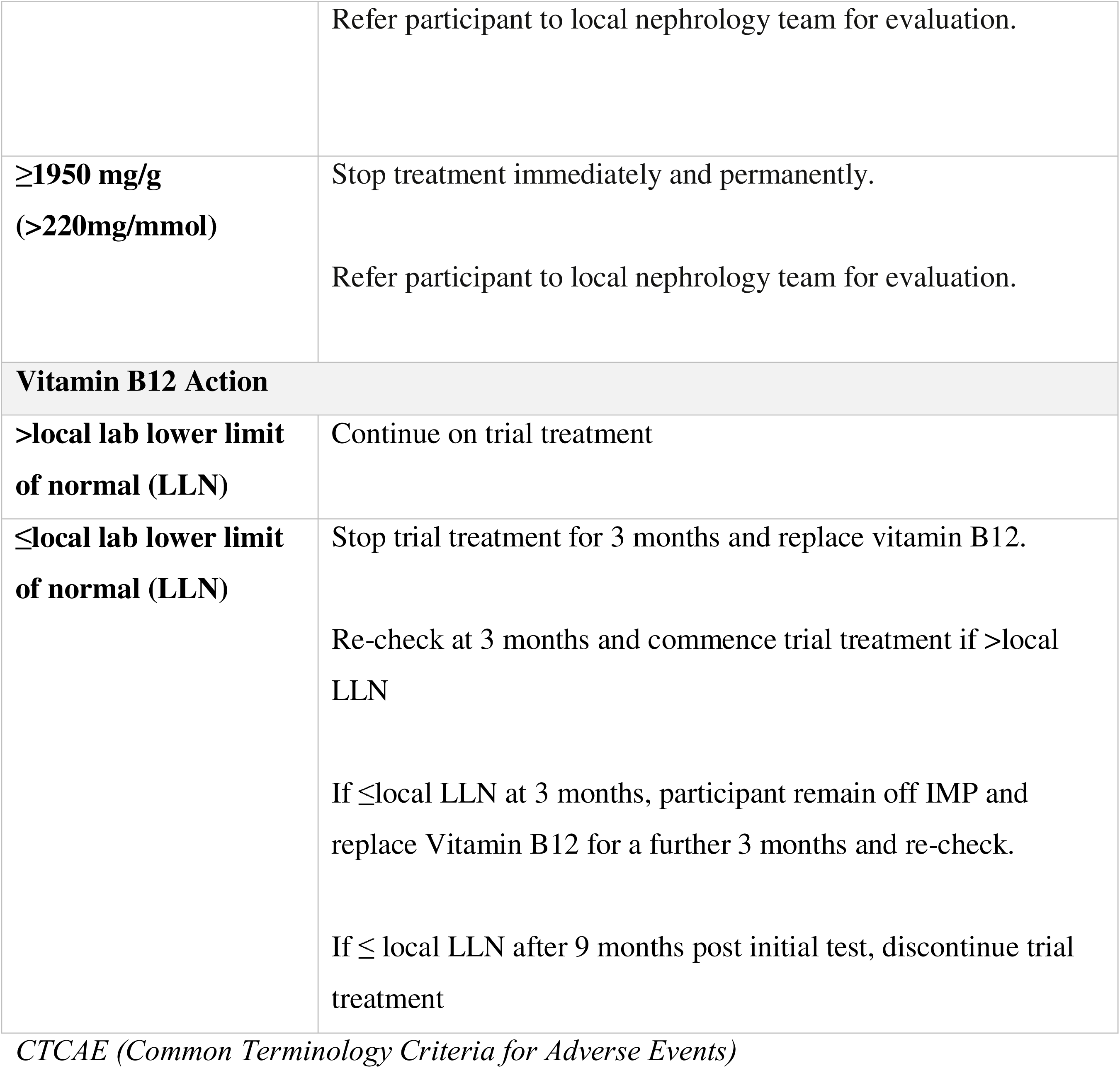
Expected Toxicities and Dose Modification/Stopping.

| **Gastrointestinal** |  |
| --- | --- |
| **CTCAE (v5.0)**<br>**Grade 1** | If on high dose, clinician discretion to consider switching to low dose, until stable<br>If on low dose, clinician discretion to consider 1 week treatment pause until stable |
| **CTCAE (v5.0)**<br>**Grade 2 or higher** | Reduce to low dose or pause (if on low dose); re-attempt dose escalation after minimum of 1 week if symptoms improve, aiming to continue at the high dose<br>If grade 2 toxicity persists, consider the following:<br>▪ Pause treatment for 2 weeks, and re-start<br>▪ And re-attempt a dose escalation at clinical discretion if toxicities are ≤ grade 1 if within 26 weeks of randomisation. |
| **eGFR** |  |
| $\geq 60$ ml/min/1.73m<sup>2</sup> | Continue on trial treatment |
| 45-59 ml/min/1.73m<sup>2</sup> | Repeat eGFR 4 weeks post-test and continue on current dose.<br>On repeat eGFR, result remains 45-59 ml/min/1.73m<sup>2</sup>;<br>Treatment must be permanently stopped |
| $<45$ ml/min/1.73m<sup>2</sup> | Treatment must be permanently stopped |
| **Proteinuria** |  |
| $\leq 300$ mg/g<br>($<34$mg/mmol) | No action required – continue on current dose |
| 301 – 1950 mg/g<br>($\geq 34$ mg/mmol – 220 mg/mmol) | Continue at current dose and retest within 2 weeks. If ACR is raised following retest ($\geq 34$mg/mmol) immediately discontinue treatment permanently. |
|  | Refer participant to local nephrology team for evaluation. |
| **≥1950 mg/g**<br>**(>220mg/mmol)** | Stop treatment immediately and permanently.<br>Refer participant to local nephrology team for evaluation. |
| **Vitamin B12 Action** |  |
| **>local lab lower limit**<br>**of normal (LLN)** | Continue on trial treatment |
| **≤local lab lower limit**<br>**of normal (LLN)** | Stop trial treatment for 3 months and replace vitamin B12.<br>Re-check at 3 months and commence trial treatment if >local LLN<br>If ≤local LLN at 3 months, participant remain off IMP and replace Vitamin B12 for a further 3 months and re-check.<br>If ≤ local LLN after 9 months post initial test, discontinue trial treatment |

*CTCAE (Common Terminology Criteria for Adverse Events)*

## OUTCOMES

### Primary outcome

The primary trial objective, achievable only on conclusion of analysis stage 2, will be to compare the effect of either active trial drug versus placebo on development of ≥6 month CDP, assessed at 6-monthly intervals, in participants with PMS based on the EDSS-Plus – a composite change in either EDSS score, and/or T25FW, and/or 9HPT compared to baseline. Progression of disability will be defined as either (i) an increase of at least 1 point if the baseline EDSS score <6, or an increase of 0.5 point if EDSS score ≥6, (ii) a 20% increase in T25FW speed (if ambulant) or subsequent inability to complete the T25FW, or (iii) a 20% increase in reciprocal combined mean time to complete the 9HPT or subsequent inability to complete the 9HPT^52^. The initial disability progression event is finalised as positive if disability is sustained and confirmed ≥6 months later in the same element of the composite, allowing for disability confirmation to take place if a subsequent visit is missed. The date of progression (if confirmed) will be the initial date of the progression event. EDSS will generally be assessed in person, but there will be capacity for this to be collected remotely via telephone ^53–55^. Time to disability progression will be compared between any of the active drug arms individually, and placebo arm, over treatment of up to 260 weeks.

To reach this point in the trial, an active treatment must pass through analysis stage 1, in which a reduction in the whole-brain atrophy rate, as measured by the SIENA technique, must be observed compared to placebo.

### Secondary outcomes

Clinician reported secondary outcomes will include: EDSS, T25FW, 9HPT, Symbol Digit Modalities Test (SDMT); Sloan Low Contrast Visual Acuity (SLCVA), MS Functional Composite (MSFC) Z score (comprising T25FW, 9HPT, and SDMT), and relapse rate.

Patient-reported outcomes will include the: Multiple Sclerosis Impact Scale-29 v2 (MSIS-29v2); Multiple Sclerosis Walking Scale-12 v2 (MSWS-12v2); Modified Fatigue Impact Scale - 21 (MFIS-21); Chalder Fatigue Questionnaire (CFQ); and Neuropathic Pain Scale (NPS).

### Health economic outcomes

For health economic analysis, the EQ-5D-5L Health Questionnaire and Client Services Receipt Inventory (CSRI) will be utilised.

### Trial schedule

Supplementary Table 1 presents the trial schedule of assessments. Following baseline and randomisation, when a full set of clinical assessments and patient-reported questionnaires will be completed, participants will attend the clinic for safety monitoring and dose escalation at 4 weeks, followed by a further visit for safety monitoring at 12 weeks. After their subsequent 26-week visit, participants will have further assessment visits every 26 weeks post-randomisation. Where participants are unable to attend their 26-weekly follow-ups, a telephone-EDSS assessment will be undertaken ^53–56^, together with remotely collected patient-reported outcomes and locally performed safety bloods.

Both the clinician-reported outcomes: EDSS, 9HPT, T25FW, SDMT, and SLCVA, and patient-reported outcomes: MSIS-29v2, MSWS-12v2, CFQ, MFIS-21, EQ-5D-5L and CSRI, will be completed every 26 weeks.

### Power and sample size

For the analysis stage 1 interim outcome assessment of PBVC to achieve 95% power to detect a 0.15% per year difference at a 1-sided level (alpha) of 35%, while allowing for a 11% drop-out, 125 participants per arm will be needed. Calculations assume an atrophy rate of 0.55% per year in the placebo arm and standard deviation of 0.55% per year at week 78, based on data from previous trials ^38,57–61^.

For the analysis stage 2 final outcome of 6-month EDSS-Plus CDP, in order to achieve 90% power to detect a hazard ratio of 0.75 (25% relative reduction) at the conventional 2-sided significance level of 5% (or 2.5% 1-sided significance equivalent), 281 progression events will be required in the placebo arm, and therefore 600 participants per arm required. This will be achieved by recruiting 125 participants to analysis stage 1 (who undergo MRI), followed by a further 475 participants to analysis stage 2 (who do not undergo MRI), for a total of 600 participants per arm. Individual participant trial duration is expected to be between 78 and 260 weeks. With 600 participants per arm it is expected that 281 progression events will be observed in the placebo arm by around 78 weeks after the last participant has been enrolled, assuming a 11% drop-out rate and 50% disability progression rate by 3 years, with the projected recruitment rate of 20 participants per month during analysis stage 1 and 40 per month in analysis stage 2, and at least one active trial arm continuing into analysis stage 2.

## ANALYSES

### Statistical analysis

The primary statistical analysis will focus on the ‘treatment policy’ estimand (an intent-to-treat analysis), with the impact of non-compliance assessed in a pre-specified sensitivity analysis targeting the ‘hypothetical’ estimand, in which the treatment effect in the absence of non-compliance is estimated (see Compliance below). In all analyses, participants in an active arm will only be compared to placebo participants with the same ‘randomisation availability’ (in the event of participants who were ineligible for one of the arms and therefore could only be randomised between two arms, a single active arm versus placebo).

As each treatment will be compared against the placebo arm only, and that the selected drugs are from different mechanistic classes, we will not adjust the type I error rate for multiple comparisons ^62,63^. Allowance for multiple comparisons will be considered if a later addition to the platform includes drugs of similar mechanistic action (e.g different doses or treatment combinations of drugs being used in other arms).

A detailed statistical analysis plan (SAP v1.0) has been written and signed off. In all stage 2 analyses, statistical tests will use a 2-sided p-value of 0.05, while the interim stage 1 analysis will use a one-sided p-value of 0.35, unless otherwise specified. Statistical analysis will be performed using Stata (StataCorp, College Station, TX, USA). Other statistical software may also be used where Stata® does not provide the relevant statistical method.

### Analysis Stage 1 (Interim) Primary Outcome

For analysis stage 1 of R/S-ALA and metformin treatments, PBVC will be assessed as a continuous variable, with pairwise comparisons between each experimental arm and placebo. This will use atrophy data measured between pairs of MRI scans taken at randomisation, week 26 and week 78 for all participants, and additionally from MRI at 104 weeks for the participants who have reached this visit (predicted to be approximately 50% of participants at the time of stage 1 analysis). The arms will be compared on the rate of change in PBVC from baseline. In the event of pseudoatrophy being evident between baseline and week 26, the arms will be compared on the rate of change from week 26 onwards.

Mean rates of PBVC will be analysed using linear mixed-effects models of repeated direct measures of change ^64^ with adjustment for the minimisation variables. All participants with at least one measure of atrophy (i.e. have at least one follow-up scan) will be included. To estimate the effect of treatment on the rate of change, an interaction will be fitted between the treatment group and the duration between each pair of MRI scans. Interactions will also be included between duration and the minimisation variables: site type (three levels), sex (male/female), age (<45, ≥45 years), baseline EDSS (4.0-5.5, 6-6.5 and >6.5), use of current DMT at baseline (yes, no), and PMS phenotype (PPMS, SPMS). Random effects will be used to account for intra-individual PBVC correlations.

A one-sided hypothesis test will be conducted, and estimated treatment effects will be reported with confidence intervals corresponding to the 0.35 alpha level in the stage 1 analysis. Based on previous trial data on PBVC^15^, it is anticipated that assumptions for the model will be met, however they will be assessed and non-parametric methods of inference will be used in the event of material violation. As there is some uncertainty around the assumption about standard deviation, a review of emerging data from the placebo arm is planned, at which point the sample size calculation will be reviewed and the alpha level may be adjusted by the TMG in agreement with the Trial Steering Committee (TSC) and Independent Data Monitoring Committee (IDMC). Similarly, a regular review of the scientific landscape will be used to ensure that the choice and structure of the interim outcome remains appropriate, with the opportunity to update it throughout the trial’s duration. These measures will ensure that 95% power can be achieved with the planned sample size.

### Analysis Stage 2 (Final) Primary Outcome

The primary outcome for analysis stage 2 will be assessed as a time-to-event variable, measuring time from randomisation to composite EDSS-Plus CDP (consisting of EDSS, T25FW, and 9HPT) initial disability progression (if confirmed ≥6 months later) between the active and placebo arms. Pairwise hazard ratios and 95% confidence intervals will be calculated using a Cox proportional hazards model, and the cumulative incidence functions over time will be produced for each treatment group. The time scale used for survival analysis will be time since randomisation. Observations will be censored for participants without an event, including those who will have withdrawn from the trial, been unable to continue, been lost to follow-up or have died due to causes other than MS (on the date of the final/most recent follow-up visit or date of death). Death due to MS will count as a confirmed progression event, as this would be an EDSS score of 10.

The model will include stratification by site type (three levels) and adjusted for the minimisation variables as fixed effects: sex (male/female), age (<45, ≥45 years), baseline EDSS (4.0-5.5, 6-6.5 and >6.5), use of current DMT at baseline (yes, no), and PMS phenotype (PPMS, SPMS)). In the event of deviation from the proportional hazards assumption, flexible parametric models will be fitted with suitable model-based survival (including difference) and hazard functions presented for illustrative purposes.

### Analysis Stage 2 (Final) Secondary Outcomes

Secondary clinician and patient-reported outcome measures for analysis stage 2 will include: EDSS, T25FW, 9HPT, SDMT, MSFC Z-score, SLCVA, and relapse count; and MSIS29v2, MSWSv2, MFIS-21, CFQ, and NPS changes, respectively. Health-related quality of life and resource use measures will include the EQ-5D-5L Health Questionnaire and Client Service Receipt Inventory (CSRI).

The rate of change in continuous clinician- and patient-reported outcomes will be compared between groups using linear mixed models for repeated-measures. For each outcome, the difference in the mean rate of change in the outcome will be reported, alongside the 95% confidence intervals and the two-sided p-value. The model will include as fixed effects (as both an intercept shift and slope interaction effect with time) site type (three levels), sex – (male/female), age (<45, ≥45), baseline EDSS (4.0-5.5, 6-6.5 and >6.5), use of current DMT at baseline (yes, no), and PMS phenotype (PPMS, SPMS).

All deaths will be dealt with using a ‘while alive’ estimand approach. With regards to the T25FW, 9HPT, SDMT, MSFC Z-score, and SLCVA, if there are missing data due to MS severity a composite approach will be used by singly imputing a relevant value. Other missing outcomes, whether due to MS severity or otherwise, will not be imputed and assumed missing-at-random.

The number of relapses in the analysis time period will be used to calculate a rate. The targeted treatment effect will be the incidence-rate ratio parameter between treatment groups using a negative binomial model (to account for expected over-dispersion where the mean does not equal the variance) of the relapse count given the analysis time (entered as an offset). The model will include as fixed effects the randomisation minimisation variables: site type (3 levels), sex – (male/female), age (<45, ≥45), baseline EDSS (4.0-5.5, 6-6.5 and ≥7.0), use of current DMT at baseline (yes, no), and PMS phenotype (PPMS, SPMS). All deaths will be dealt with using ‘while alive’ estimand approach. However, death due to MS will also count as an additional relapse.

### Compliance

Compliance will be assessed from the dose each participant is on during the trial and the number of days participants missed taking IMP over the 30 day periods following each trial visit. The dose given to each participant, including any dose changes, will be recorded in the trial database. Participants will be provided with a drug diary to record their uptake of trial medication between clinic visits. The diary card will be returned at each visit, and participants will be invited to complete a summary of missed doses on the electronic system directly or this will be entered by site staff from the returned diary card.

Participants will be considered compliant with their randomised intervention if they were on the per-protocol dose for at least 90% of days during the trial and took all their prescribed IMP on at least 90% of days, based on self-reported medication use.

### Missing data

Missing data will be identified, and efforts will be made to obtain them from the original source medical records. Characteristics of participants who withdrew and reasons for withdrawal will be tabulated by treatment group and compared with those with complete data in the IDMC report. In the event of substantial differences in withdrawal patterns being found, further sensitivity analyses will be carried out to assess the robustness of the results. For analysis stage 1 MRI brain atrophy assessment, missing data will not be imputed, with the assumption that the reasons for missingness are non-informative. For stage 2 analysis, data will only be used for visits where at least one of the EDSS, T25FW or 9HPT is recorded (including singly imputed values for T25FW and 9HPT for missing values due to MS severity). It is assumed that interval censoring of missing values conveys no further information, and that censoring withdrawing patients is non-informative with the same progression rate as those who continue to be followed up.

### Health economic analysis

If effective, the very low cost of these IMPs means there is a high probability that they will be cost-effective compared to current practice, due to potential reductions in other resource use. Resource use will be collected using a patient resource use measure (RUM) modified to capture PMS-relevant resource use (collected at baseline and 6-monthly intervals), and serious adverse events (SAEs) recorded as part of the trial. The RUM will include primary and MS specialist secondary care, paid and unpaid carer time, complementary therapies and work absenteeism. Resource use will be obtained using published sources, including the National Cost Collection for the NHS, Unit Costs of Health and Social Care^65^ and the British National Formulary^66^. The difference in costs between arms will be calculated using ordinary least squares regression, accounting for stratification by site and adjusting for baseline costs and minimisation factors as defined in the statistical analysis, with 95% confidence intervals generated using bias-corrected and accelerated bootstrap estimations. We will conduct sensitivity analyses, calculating the incremental cost by arm using general linear models. The primary analysis will be from an NHS (UK) and Medicare (AUS) and personal social services perspective, with secondary analyses from wider societal perspectives.

The mean incremental cost per quality-adjusted life year (QALY) will be estimated using the EQ-5D-5L (collected at baseline and 6-monthly intervals) to calculate the utility score at each time point, using the algorithm recommended by NICE and analysed using an area under the curve approach ^67,68^. Furthermore, given uncertainties regarding the suitability of the EQ-5D-5L for people with PMS ^67^, an MS-specific measure – the MSIS-29v2^69^ – will be considered for estimating QALYs using methods available in the literature ^70^. The difference in QALYs between arms will be calculated using ordinary least squares regression, accounting for stratification by site and adjusting for baseline utilities and minimisation factors as defined in the statistical analysis. 95% confidence intervals will be generated using bias-corrected and accelerated bootstrap estimations. The primary analysis will be using the EQ-5D-5L to calculate QALYs, with a secondary analysis using the MSIS-29v2.

Two analysis approaches will be utilised: (i) ‘within trial’ period patient level data, and (ii) a model-based approach for the lifetime horizon of the patient. In the case of the latter, a Markov model using EDSS states will be used to model expectations beyond the trial duration. Both analyses will include a secondary wider cost perspective including: the cost of the impact on carers, absenteeism and out-of-pocket costs.

The within-trial economic evaluation will be conducted using a cost-effectiveness estimate, comparing the intervention to control over the trial period to provide an incremental cost-effectiveness ratio (ICER). Confidence intervals for QALYs and mean costs will be calculated using a non-parametric bootstrap with replacement, presented on a cost-effectiveness plane, and a cost-effectiveness acceptability curve will be constructed. The probability that the intervention is cost-effective for different QALY value thresholds will be determined from the latter. Missing trial data will be handled using appropriate methods such as multiple imputation. A detailed economic evaluation analysis plan will describe the methods and be presented to the TSC for approval. Differences in follow-up duration will be accounted for by including exposure duration in the analysis.

An incremental cost-effectiveness ratio (ICER) will also be reported from a model-based analysis; it will be based primarily on the trial data alongside published sources, modelling QALYs and predicted costs according to EDSS states using a Markov model. This will allow progression to be simulated through different health states over time and estimations of changes in health-related quality of life and costs to be extracted.

Good practice guidelines for economic evaluations will be used for the analysis^70^. Long term costs and health outcomes will be discounted using discount rates recommended by NICE^71^.

## MONITORING

### Oversight

The Trial Sponsor (UCL), with delegated authority to the Innovative Clinical Trials Unit (InCTU) at UCL (formerly the Medical Research Council Clinical Trials Unit) is formally responsible for the oversight of the trial, ensuring it is conducted in compliance with Good Clinical Practice (GCP), relevant regulations and ethics committee permissions. A Trial Steering Committee (TSC) provides overall supervision for the trial and provides advise through its independent chair. The Independent Data Monitoring Committee (IDMC) will independently review and consider unblinded data to advise the TSC. The Trial Management Group (TMG) will be responsible for the trial execution and oversight. Following advice by the IDMC, the ultimate decision for trial/arm continuation or closure will lie with the TSC in consultation with the TMG. The full structure of the trial committees and description of their responsibilities are highlighted in Supplementary Figure 1.

### Safety event reporting

Site investigators will report all adverse events and reactions (AEs and ARs) irrespective of severity, causality or expectedness in relation to the trial intervention, which take place from randomisation to the end of patient pathway (including 4 weeks after discontinuation of trial treatment) to the InCTU at UCL via the electronic data capture (eDC) system within 1 week of notification. Adverse events and reactions meeting the ‘serious threshold (SAEs and SUSARs) will be subject to expedited reporting to the Sponsor (immediately and no later than 24 hours from site awareness) in line with the relevant UK and Australian regulations.

Adverse events will be recorded and graded according to the Common Terminology Criteria for Adverse Events (CTCAE) v5.0, using a recognised medical term or diagnosis that accurately reflects the event. Adverse events will be assessed by the local investigator for severity, relationship to the investigational product, possible aetiologies, and whether the event meets criteria of an SAE and therefore requires expedited notification.

Medically qualified staff in the OCTOPUS Trial Management Team (TMT) at the InCTU at UCL will review all SAE reports received, code to Medical Dictionary for Regulatory Activities (MedDRA) and perform the expectedness assessment using the approved Reference Safety Information (RSI). They will then report them to the Medicines and Healthcare products Regulatory Agency (MHRA) and the research ethics committees, as appropriate, and as part of Annual Safety Reports. The TMT will also keep all investigators informed of any safety issues that arise during the trial.

Other notable events that require expedited reporting to the OCTOPUS TMT at InCTU at UCL include lactic acidosis, glomerulonephritis, microalbuminuria, and macroalbuminuria, which are toxicities of interest for the current OCTOPUS IMPs, and as such will be reported via the eDC system within the same timelines as for SAEs.

Additionally, though pregnancy is not an adverse event, if a pregnancy occurs in a trial participant or a partner of a trial participant, it is a notable event and female trial participants must stop trial treatment. Again, the sponsor will be notified within 24 hours of the site becoming aware of the event and with consent, all pregnancies will be followed up to collect information until 30 days following the outcome of the pregnancy, end of treatment period or end of trial regardless of the outcome.

### Audit

Data quality control and assurance will be conducted via both on-site monitoring and central monitoring at the InCTU at UCL. The frequency, type and intensity of routine monitoring and the requirement for triggered monitoring is decided by the Quality Management and Monitoring Plan (QMMP). On-site monitoring will be adaptive and live, occurring as data are entered into the eDC^72,73^. Central data review will then audit data entered into the OCTOPUS eDC system for errors, missing data points and protocol deviations and will raise queries as appropriate.

### Emergency contacts and unblinding

Unblinding of allocation to trial treatment can be performed, if required, only in a medical emergency where knowledge of the participant’s treatment allocation would change clinical management. Unblinding can be performed by any treating doctor, the participant’s trial site PI, delegated site clinicians, OCTOPUS CI or Trial Physicians via the OCTOPUS website 24 hours, 7 days a week.

All recruited participants will be given a Participant’s Card with the details of the OCTOPUS website, where the unblinding can be performed, and contact details of the participant’s site clinical trial team. The trial site PI, delegated site clinicians, OCTOPUS CI or Trial Physicians will receive notification that the participant has been unblinded but will not receive details of treatment allocation. This must then be reported to the OCTOPUS trial team within 24 hours.

### Withdrawal of trial participants

Participants may withdraw from trial participation at any time without prejudicing their right to standard of care. Participants who discontinue the trial intervention will remain in follow-up unless they withdraw consent. Data from withdrawn participants, up to the date of their withdrawal, will be included in the analysis unless they withdraw consent for all data being held. Participants who stop trial follow-up early will not be replaced.

### Patient and Public Involvement and Engagement

Since the earliest study concept meetings in 2018, the OCTOPUS trial has had a strong PPIE strategy, with significant contributions from the UK MS Society’s Research Network and building on related broader efforts to enhance inclusivity in MS clinical trials ^74^, with a PPIE member (SS) having been a co-applicant on the original award and acting as a member of the subsequent TMG.

The strategy was shaped by discussions at numerous workshops across the UK, attended by people with MS, which considered various topics, including but not limited to recruitment information methods, clarity of trial information, entry criteria (age and disability ranges), incentives and barriers to participation, and outcome measures.

People affected by MS will remain embedded in the trial’s operational functioning, with active involvement in the TMG, Treatment Advisory Committee (TAC), TSC, and various TMG subgroups. There will also be a dedicated PPIE Forum for OCTOPUS set up by the UK MS Society. All PPIE members will be supported by the MS Society Public Involvement Manager and a member of the OCTOPUS team at the INCTU at UCL.

PPIE input via the UK MS Society PPIE Forum will shape the format, content and timing of trial updates. This will ensure they are appropriately worded, facilitate retention, and play a key role in communicating the results of the study. We will also seek feedback from OCTOPUS participants on their experiences of the trial, to gain an understanding of what could be done differently in future.

## ETHICS AND DISSEMINATION

### Data management and confidentiality

All trial documentation (consent forms, worksheets (where used as source data for Electronic Case Report Form, eCRF), clinical notes and administrative documents) will be kept in secure, restricted access locations archived for a minimum of 25 years after the end of the trial, during which it will be accessible to the competent or equivalent authorities, the Sponsor, InCTU at UCL and other relevant parties in accordance with the applicable regulations. Where documentation is in the form of electronic health records, these will be stored in accordance with local guidelines and retained for the maximum period permitted locally.

Trial data will be entered by trained site personnel into a secure, validated online database (Open Clinica), an eDC system used by the MRC Clinical Trials Unit at UCL, accessible only by delegated team members at that site, and by delegated staff from the TMT. Such records should include only the nine-digit *Participant identification number* and a *three-letter code* (TLC) – allocated at time of trial consent – to identify the participant. Under the UK Data Protection Act 2018 and Australian Privacy Act 1988, the latter identifiers will be considered as sensitive personal data and will be treated as such by both the site team and the TMT.

As well as data entered by site personnel, participants will also be able to enter certain data directly into the eDC, including all patient-reported outcome measures (except the Client Services Receipt Inventory, CSRI) and Drug Diary Cards.

### Patient consent

All OCTOPUS participants will need to provide informed consent prior to any trial-specific procedures being undertaken in compliance with the approved protocol, the Declaration of Helsinki 1996, the principles of Good Clinical Practice (GCP) as laid down by the ICH topic E6 (R2), Commission Clinical Trials Directive 2005/28/EC*. Implementation in national legislation in the UK is by Statutory Instrument 2004/1031 and subsequent amendments, the UK Data Protection Act 2018 (DPA number: Z6364106), and the UK Policy Framework for Health and Social Care Research. Implementation in national legislation in Australia is by the Privacy Act 1988 (Cth), Australian Privacy Principles, National Statement on Ethnical Conduct in Human Research, Australian Clinical Trial Handbook (2024), Therapeutic Goods Act 1989, Australian Code for Responsible Conduct of Research, and National Operating Procedures for Clinical Trials. Participants will also agree to Sponsor review of their medical records for clinical trial monitoring or inspection purposes.

### Ethical approval and dissemination

The National Research Ethics Service Committee (London, Hampstead) reviewed the trial protocol and materials to be given to participants (approved 14^th^ October 2022, REC ref 22/LO/0622). The trial protocol and materials to be given to Australian participants has also been reviewed and approved by the Gold Coast Hospital and Health Service, Human Research Ethics Committee (approved 16^th^ December 2024, HREC/2024/QGC/97381). This article refers to the current protocol (Version: 8.0, Date: 28^th^ August 2025). The findings of this trial will be disseminated through peer-reviewed publications and conference presentations. There will be a close communication strategy developed with the UK MS Society and full PPIE engagement.

### Access to data

Data will be stored on an encrypted and password-protected database. Site staff will have direct control of their own site’s data. Where future analyses are proposed, these will be considered by the TMT, TMG and TSC, with appropriate data-sharing formal agreements to be put in place.

## DISCUSSION

The OCTOPUS trial represents a significant step forward in charity-funded investigator-driven clinical trials for slowing Progressive Multiple Sclerosis which is the dominant unmet need for this condition. Trial development has been the culmination of the organisational, supportive, and financial efforts of the MS Society and researchers to expand traditional methods, such as the ealier single cycle/stage multi-arm MS-SMART trial. OCTOPUS has been developed with contributions by a large team of people with lived-experience, experts in trial design from the InCTU UCL, experimental and translational scientists, neurologists, pathologists, radiologists, and pharmacologists. It utilises efficient adaptive platform trial methodologies and state of the art statistical approaches to deliver clinically meaningful answers with greater speed and inclusivity at lower costs. The trial has been funded by the peak bodies for MS research support in the UK and Australia, and provides an opportunity for a significant number of people with PMS to engage in ground-breaking research.

This protocol publication is based on version 8.0. Since the trial launch, there have been five significant changes of note. Monitoring of eGFR cut-offs was updated to reflect clinical practice with metformin^75^ in version 4.0, with monitoring for eGFR values between 45-60 mL/min/1.73m^2^ and treatment cessation if the eGFR reduced below 45mL/min/1.73m^2^. Version 5.0 was updated to include details and procedures relevant to the Australian sites and specific to discontinue trial medication if an ACR value >113mg/mmol is found prior to repeat testing. Version 8.0 included an update to the screening and monitoring of proteinuria using ACR rather than solely urine dipsticks. A full list of protocol updates is included in the supplementary material. Additionally, a further 4 sites across the UK are being set up as part of analysis stage 2, and the Australian extension, Platform Adaptive Trial for Remyelination and Neuroprotection in Multiple Sclerosis (PLATYPUS), has launched, with a further 9 sites currently in set-up.

The trial launched in January 2023 and concluded stage 1 recruitment of 384 participants in November 2024. Analysis stage 2 recruitment is ongoing, with a total of 787 participants having been randomised to the trial as of the 1^st^ June 2026 across 24 active sites in the UK, and 1 site in Australia. Analysis stage 1 interim outcome assessment is planned for late 2026.

## Data Availability

N/A - no data is presented in the current work as it is a protocol paper

## ACKNOWLEDGEMENTS

Foremost, we thank the participants in this trial for their tireless support, dedication and commitment. We thank all individuals who contributed to a full, continuing and meaningful PPIE. Our thanks to Dr Emma Gray and colleagues at the UK MS Society for their complete and on-going support of OCTOPUS. We acknowledge the contributions of all members of the Treatment Selection Group, whose work laid the foundation for the Treatment Advisory Committee’s activities. We thank JCampbell for further oversight and conduct of the Australian health-economic evaluation in PLATYPUS. We also acknowledge the support of the independent members of the Trial Steering Committee (TSC), the Independent Data Monitoring Committee (IDMC), and the MS Society’s OCTOPUS Oversight and Management Committee (OMC). Finally, we thank all OCTOPUS Investigators and PLATYPUS Investigators.

## CONTRIBUTORS

JChataway, MP, FB, AJT, OC, SC, SP, DF, MB, JN, SS, RM, and ET researched and wrote the original grant application. All authors are involved in the setup and running of the trial and contributed to the writing of the manuscript. CP represents the sponsor and developed and oversees the trial protocol and processes. SAM and CW led the writing of this manuscript. MBurnell, JN, RB, JCarpenter, MP, RH, and EB were involved in the development of the statistical and health economic analysis facets of the trial, protocol design and contributed to the writing of the manuscript. SAM, CW, ET and PF provide clinical and safety review support to the trial. SAM, CW and MBraisher comprised the lead site clinical team working on the trial and contributed to the writing of the manuscript. CP, AN, EB, and ML are the trial management team at the UCL InCTU and contributed to the writing of the manuscript. HS, MR and CP programmed the database and set up data management processes and systems. SS is the PPIE representative and critically reviewed the manuscript. RA-F, TA, PF, HLF, IG, JG, CH, MM, GM, OP, SP, CR, BS, SAB, BY, ES, CS, AD, ET and JChataway are site principal investigators and contributed to the writing of the manuscript. AW and DF led the TAC advice for selection of trial drugs and contributed to the writing of this manuscript.

JChataway is the chief investigator for the trial and contributed to the protocol and the manuscript. JChataway is the guarantor.

## COMPETING INTERESTS

### Declarations

In the last 3 years, JChataway has received support from the Health Technology Assessment (HTA) Programme (National Institute for Health and Care Research, NIHR), the UK MS Society, and the US National MS Society and the Rosetrees Trust. He is supported in part by the National Institute for Health and Care Research, University College London Hospitals (UCLH) Biomedical Research Centre, London, UK. He has been a local principal investigator for a trial in MS funded by MS Canada. A local principal investigator for commercial trials funded by: Ionis and Roche; and has taken part in advisory boards/consultancy for: Biogen, Contineum Therapeutics, FSD Pharma, InnoCare, Pheno Therapeutics and Roche.

In the last 5 years ET has received honorarium for consulting work from Biogen, Janssen, Merck, Novartis, and Roche. She has received travel grants to attend or speak at educational meetings from Biogen, Merck, Neuroax, Roche, and Novartis.

MP declares institutional educational grant funds or support for medicines from Novartis, Pfizer, Roche, Roche Products, Sanofi, Serum Institute of India, Shionogi, SUMVAX, Synteny Biotechnology, Takeda, Tibotec, Transgene, ViiV Healthcare, Virco Xenothera, CSL Behring, Eli-Lilly, Emergent Biosolutions, Gilead Sciences, GlaxoSmithKline, Grifols, ICON, Janssen Products, Janssen-Cilag, Janssen Pharmaceutica, Johnson & Johnson, Merck Serono, Micronoma, Modus Theraputics, MSD, Mylan, Abcodia, Advanced Accelerator Applications International, Akagera Amgen Aspirin Foundation, Astellas, AstraZeneca, AoA, Baxter, Bayer, Bristol Myers Squibb US, Bri-Bio B&C Group, Cepheid, Cipla, and Clovis

JCarpenter reports grants from UK Medical Research Council, personal fees from Wiley, personal fees from Springer, personal fees from University of Bern, personal fees from Statisticians in the Pharmaceutical Industry, and personal fees from Novartis.

RH received grants from NIHR DemPRU-QM, NIHR Evidence Synthesis, and NIHR Health Protection Research Unit; received consulting fees from Ministry of Justice, UK Health Security Agency, University of Nottingham, and QuidelOrtho; and leadership role on transforming health and care systems EU funding board as Chair of the Board.

AJT receives a fee from being Co-Chair, University College London (UCL)-Eisai Steering Committee drug discovery collaboration; German Aerospace Center, Heath Research (ERA-NET NEURON); consultancy from Sandoz Global Advisory and Novartis. Member, National Multiple Sclerosis Society (USA) Research Programs Advisory Committee; Clinical Trials Committee, Progressive MS Alliance; Board member, European Charcot Foundation; Editor in Chief, Multiple Sclerosis Journal; Editorial Board Member, The Lancet Neurology. AJT has received fees for academic reviews for Jockey Club College at City University of Hong Kong, Health Research Board Ireland, Sant Pau Biomedical Research Institute. AJT has received support from the UCL/ UCLH NIHR Biomedical Research Centre. AJT received travel support from European Committee for Treatment and Research in Multiple Sclerosis (ECTRIMS), the European Charcot Foundation, Health Research Board Emerging Clinical Scientist Award, Polish Neurological Society, and Armin Curt Farewell Symposium. He receives no fee from being Chair (Scientific Ambassadors), Stop MS Appeal Board, UK MS Society; Research and Academic Counsellor, Fundació Privada Cemcat; Ambassador, European Brain Council. AJT additionally holds a patent for the MSIS-29 Impact Scale.

FB is supported by the NIHR biomedical research centre at UCLH. He has been: on the steering committee or Data Safety Monitoring Board member for Biogen, Merck, Eisai, Prothena and Idorsia; advisory board member for Combinostics, Scottish Brain Sciences, IXICO and Alzheimer Europe; consultant for Roche, Celltrion, Merck, Bracco; research agreements with ADDI, Merck, Biogen, GE Healthcare, Icometrix, Roche; co-founder and shareholder of Queen Square Analytics LTD

OC declares being NIHR Research Professor (RP-2017-08-ST2-004); over the past 2 years, member of independent data safety and monitoring board for Novartis; has received consulting fees or speaker honoraria from Lundbeck, Merck or Biogen; contributed to an advisory board for Biogen; she is Deputy Editor of Neurology, for which she receives an honorarium; has received research grant support from the UK MS Society, the NIHR UCLH Biomedical Research Centre, and the NIHR; and is vice-president of ECTRIMS (unpaid).

EG is Director of Research at the UK MS Society and has no declarations.

CR has received research grants from the UK Medical Research Council, Sanofi, Burden Neurological Society, University of Bristol, and Bristol Health Research Charity, and support for clinical research for the OCTOPUS trial (UK MS Society). CR has participated on a data safety monitoring board for the CCMR Two and NEuEoMS trials. Unpaid advisory roles with the UK MS Society, Burden Neurological Institute, Association of British Neurologists, and NICE HTA Assessment Group.

HLF has received honoraria for advisory boards or educational activities from Merck, Novartis, and Roche. HLF has research grant support from the NIHR Health Technology Assessment Programme and Efficacy and Mechanisms Evaluation, UK MS Society, and the Horne Family Charitable Trust. She has been a local PI for multiple sclerosis clinical trials of an investigational medicinal product funded by Novartis, Roche, and Biogen Idec. LF has received honoraria for speaking or as an advisory board member from Biogen, Novartis, Merck, Sanofi Genzyme, and Roche and received support for attending educational meetings from Biogen, Novartis, Merck, Sanofi Genzyme, and Teva.

MM has received travel support, speaker honoraria, or consultation fees from Merck-Sereno, Novartis, and Roche. RN receives support from UK MS Society to Imperial College and Swansea university; is a trustee of the Multiple Sclerosis Trials Collaboration charity; and attended paid advisory boards for Novartis and Roche.

IG received the following funding in the last 36 months: research grants from Medical Research Council UK, National Institute for Health and Care Research, MS Society UK, Wessex Medical Research, Independent Research Fund Denmark, Rosetrees Trust, Guarantors of Brain, Kedrion, The Binding Site (Part of Thermo Fisher Scientific), and Evgen; speaker honoraria and travel funding from Novartis. D.G. and V.I. are employed by Binding Site (Part of Thermo Fisher Scientific).

MBurnell has been funded by grants from the Medical Research Council (MRC), Cancer Research UK, the National Institute for Health Research (NIHR), and The Eve Appeal.

RAF has participated on advisory boards, received travel grants and attended educational programmes sponsored by Roche and Novartis.

SAB has been awarded funding by the Medical Research Future Fund, MS Australia and the Trish Foundation. He is a principal investigator for a clinical trials sponsored by Novartis and Sanofi, and has received speakers fees from Novartis for chairing scientific meetings.

SAB has been a principal investigator in clinical trials sponsored by Biogen Idec, Novartis, Genzyme and ATARA Biotherapeutics.

ORP has received honoraria and travel expenses from Biogen, Bayer, Genzyme, Merck, Novartis, Roche, Sanofi and Teva and served on advisory boards/acted as a speaker for Biogen, Celgene, Janssen, Merck, Neuraxpharm, Novartis, Roche and Sanofi.

SPl is founder, chief scientific officer, and shareholder (>5%) of Cambridge Innovation Technologies Consulting and is a consultant for Aspen Therapeutics, Secretome Therapeutics, Macomics, Solute Guard Therapeutics, and Astex Pharmaceutical.

SPa has been funded by grants from the National Institute for Health and Care Research (NIHR) and is a hub lead for the NIHR Leeds Clinical Research Facility.

In the last 3 years DF has received research support from the UK MS Society, Higher Education Authority (Rep. of Ireland), Wellcome, Research Ireland and Sangamo Inc. She received consultancy fees for participation in an advisory board for Sangamo Inc.

TA has received honoraria or consulting fees for participating in advisory boards related to trial steering committees and data and safety monitoring committees, speaker fees & research grants Janssen, Merck, Novartis, Roche and Sanofi-Genzyme. She is partially funded by the NIHR SCPRA fellowship.

All other authors declare no competing interests.

