## Supplementary material for "Optimal Clinical Trials Platform for Progressive Multiple Sclerosis (OCTOPUS): protocol for an international, multi-arm, multi-stage, platform, randomized controlled, double-blind, phase 3 clinical trial"

### 1. Full list of OCTOPUS Investigators and MRC CTU at UCL Team

| **Trust name** | **Hospital** | **Town/City** | **Name** | **Role** |
| --- | --- | --- | --- | --- |
| Cambridge University Hospitals NHS Foundation Trust | Addenbrookes Hospital | Cambridge | Stefano Pluchino | Principal Investigator |
|  |  |  | Alanna Beasley | Research Nurse |
|  |  |  | Luca Peruzzotti-Jametti | Co-Investigator |
|  |  |  | Toni Bray | Research nurse |
|  |  |  | Cyrus Daruwalla | Blinded assessor |
|  |  |  | Emily Bridges | Pharmacy technician |
|  |  |  | Nicholas MacCaulliffe | Pharmacy technician |
|  |  |  | Georgina Chivers | Blinded assessor |
|  |  |  | Laura Canna | Research Nurse |
|  |  |  | Lucy Chapman | Research Nurse |
|  |  |  | Debbie Read | Research Nurse |
|  |  |  | Fiona Lee | Research Nurse |
|  |  |  | Naval Vyse | Pharmacist |
|  |  |  | Mark Bolton | Pharmacist |
|  |  |  | Ilse Patterson | Radiographer |
|  |  |  | Robyn Staples | Pharmacist |
|  |  |  | Alexander Murley | Blinded assessor |
|  |  |  | Nicholas Cunniffe | Blinded assessor |
|  |  |  | Kay Wooldridge | Pharmacy technician |
| Belfast Health and Social Care Trust | Belfast City hospital | Belfast | Gavin McDonnell | Principal Investigator |
|  |  |  | Fiona Magill | Research Nurse |
|  |  |  | Joanne Burns | Radiographer |
|  |  |  | Brian Wells | Pharmacist |
|  |  |  | Stella Hughes | Co-Investigator |
|  |  |  | Fiona Kennedy | Co-Investigator |
|  |  |  | Rachael Kee | Co-Investigator |
|  |  |  | Loreta dela Rosa | Research Nurse |
| Belfast Health and Social Care Trust | Belfast City hospital | Belfast | Aine Redfern-Walsh | Research Nurse |
|  |  |  | Karan Smyth | Health Care Assistant |
|  |  |  | Joanna Shooter | Clinical Trials Practitioner |
|  |  |  | Angelina Madden | Lab Technician |
|  |  |  | Nogol Motamedgori | Blinded assessor |
|  |  |  | Reagan O'Kane | Blinded assessor |
|  |  |  | Conor Hughes | Clinical Research Fellow |
|  |  |  | Gerry Mullan | Co-Investigator |
|  |  |  | Ben Sullivan | Blinded assessor |
|  |  |  | Sharon Carr | Research nurse |
|  |  |  | Peter Gray | Pharmacy technician |
| Bradford Teaching Hospitals NHS Foundation Trust | Bradford Royal Infirmary | Bradford | Cord Spilker | Principal Investigator |
|  |  |  | Marineo Llanaj | research nurse |
|  |  |  | Iain Hetherington | Clinical Trial assistant |
|  |  |  | Outi Quinn | Research Nurse |
| Dartford and Gravesham NHS Trust | Darent Valley Hospital | Dartford | Anisha Doshi | Principal Investigator |
|  |  |  | Angela Agore | Research nurse |
|  |  |  | Ajison Baby | Research Nurse |
|  |  |  | Guru Prasad Kumar | Co-investigator |
|  |  |  | Andrea Davis-cook | Pharmacist |
|  |  |  | Olayide Okunowo | Blinded assessor |
|  |  |  | Katrina Potter | Blinded assessor |
|  |  |  | Elaine Strachan | Clinical trials pharmacist |
|  |  |  | Kelly Clarke | Blinded assessor |
|  |  |  | Meenakshi Venkatesh | Clinical Trial assistant |
| East Suffolk and North Essex NHS Foundation Trust | Ipswich Hospital | Ipswich | Simon Kerrigan | Principal Investigator |
|  |  |  | Jenny Finch | Research Nurse |
|  |  |  | Matthew Howlett | Pharmacist |
|  |  |  | Deborah Beeby | Research Nurse |
|  |  |  | Georgina Lloyd | Data co-ordinator |
|  |  |  | Hollie Bamford | Blinded assessor |
|  |  |  | Amra Hasanovic | Pharmacist |
| East Suffolk and North Essex NHS Foundation Trust | Ipswich Hospital | Ipswich | Mortada Elyas | Co-Investigator |
|  |  |  | Clare Galton | Blinded assessor |
|  |  |  | Andrew Graham | Blinded assessor |
| The Leeds Teaching Hospitals NHS Trust | Leeds General Infirmary | Leeds | Helen Ford | Principal Investigator |
|  |  |  | Linford Fernandes | Co-Investigator |
|  |  |  | Prisca Mpofu | Research Nurse |
|  |  |  | Martin O'Malley | Research Nurse |
|  |  |  | Nathan Alldred-douglas | Senior clinical trials assistant |
|  |  |  | Helen Thorp | Pharmacist |
|  |  |  | Joanne Jackson | Pharmacist |
|  |  |  | Shanaz Begum | Research Nurse |
|  |  |  | Lucy Vinter | Research Nurse |
|  |  |  | Vinjam Maruthi | Blinded assessor |
|  |  |  | Oliver Lily | Co-Investigator |
|  |  |  | Amr Tageldin | Blinded assessor |
|  |  |  | Abdel Rahim Elniel | Blinded assessor |
|  |  |  | Gurpreet Sehmbi | Blinded assessor |
|  |  |  | Hibbah Nadeem | Clinical Research Co-ordinator |
|  |  |  | Claire Reidy | Clinical Research Co-ordinator |
|  |  |  | Sadaf Mehboob | Blinded assessor |
|  |  |  | Angela Dunne | Clinical Trial assistant |
| Swansea Bay University Health Board | Morriston Hospital | Swansea | Owen Pearson | Principal Investigator |
|  |  |  | Caroline Parsley | Research Nurse |
|  |  |  | Claire Stafford | Research Nurse |
|  |  |  | Nicola Chilcott | Research Nurse |
|  |  |  | Reece Dower | Data and Finance Assistant |
|  |  |  | Paul Jones | Pharmacist |
|  |  |  | Catherine Hughes | Pharmacy technician |
|  |  |  | Lily-Ann Peace | Research nurse |
| Mid-Yorkshire NHS Trust | Pinderfields Hospital | Wakefield | Bindu Yoga | Principal Investigator |
|  |  |  | Sarah Watson | Clinical Trial assistant |
|  |  |  | Courtney Greaves | Research Nurse |
|  |  |  | Kiren Aziz | Pharmacist |
|  |  |  | Judy Olivia Archer | Co-investigator |
|  |  |  | Dhurgham Alomar | Blinded assessor |
|  |  |  | Claire Hutsby | Pharmacy technician |
|  |  |  | Darren Gomersall | Pharmacy technician |
|  |  |  | Victoria Carley | Pharmacy support worker |
|  |  |  | Donna Exley | Research nurse |
|  |  |  | Charity Kimaru | Research nurse |
|  |  |  | Amr Tageldin | Blinded assessor |
| University Hospitals Dorset NHS Foundation Trust | Poole Hospital | Poole | Charles Hillier | Principal Investigator |
|  |  |  | Judith Dube | Research Nurse |
|  |  |  | Alison Fletcher | Radiographer |
|  |  |  | Alison Hogan | Pharmacist |
|  |  |  | Miss Julie Thomson | Data Manager |
|  |  |  | Michelle Davies | Blinded assessor |
|  |  |  | Rhiannon Smith | Blinded assessor |
|  |  |  | Nicola Hare | Blinded assessor |
| University Hospitals Dorset NHS Foundation Trust | Poole Hospital | Poole | Peter Grenholm | Co-Investigator |
|  |  |  | Chia-Hui Elaine Lim | Co-Investigator |
|  |  |  | Miss Emma Gunter | Research Nurse |
|  |  |  | Dinithi Juliyange | Co-Investigator |
|  |  |  | Luke Vamplew | research project co-ordinator |
|  |  |  | Shamayne Evetts | Blinded assessor |
|  |  |  | Brooke Hedges | Data management assistant /  Clinical Research Co-ordinator |
|  |  |  | Heidi Smith | Clinical Research Co-ordinator |
|  |  |  | Maxine Ashton | Clinical Research Co-ordinator |
|  |  |  | Sharon Power | Pharmacy clinical trials co-ordinator |
| Lewisham and Greenwich NHS Trust | Queen Elizabeth Hospital | Woolwich | Eli Silber | Principal Investigator |
|  |  |  | Eti Omoregie | Senior Clinical Trials Coordinator |
|  |  |  | Ashley Payne | Research Nurse |
|  |  |  | Georgios Dervenoulas | Blinded assessor |
|  |  |  | Liban Bussuri | Blinded assessor |
| Barking, Havering University Hospitals NHS Trust | Queens Hospital | Romford | Miriam Mattoscio | Principal Investigator |
|  |  |  | Ramya Shamji | Research Nurse |
|  |  |  | Grace Fawehinmi | Research Nurse |
|  |  |  | Mohammed-Rashid Khan | Pharmacist |
|  |  |  | Laura Azzopardi | Co-investigator |
|  |  |  | Janice Hastings | Research administrator |
|  |  |  | Parveen Dugh | Pharmacy technician |
|  |  |  | Bolanle Lawal | Clinical trials pharmacist |
|  |  |  | Laura Parker-Wall | Research Nurse |
| Barking, Havering University Hospitals NHS Trust | Queens Hospital | Romford | Elisa Visentin | Research Nurse |
|  |  |  | Renato Oliveira | Blinded assessor |
|  |  |  | Dilini Chandrasiri | Blinded assessor |
| Nottingham University Hospitals NHS Trust | Queens Medical Centre | Nottingham | Rasha Abdel-Fahim | Principal Investigator |
|  |  |  | Nichola Sherry | Research Nurse |
|  |  |  | Jerin Mary Jose | Research Nurse |
|  |  |  | Maria Scott | pharmacist |
|  |  |  | Richard Barks | Data Manager |
|  |  |  | Andrew Cooper | Radiographer |
|  |  |  | David Beesley | Pharmacy technician |
|  |  |  | Rebecca Boulton | Research nurse |
|  |  |  | Muniba Hussain | Clinical trials pharmacy technician |
|  |  |  | Karolina Beresford | Clinical Trial assistant |
|  |  |  | Ryan Mamun | Blinded assessor |
|  |  |  | Nikos Evangelou | Co-investigator |
|  |  |  | Emily Stone | Research Nurse |
|  |  |  | Grana Joice Daniel | Research Nurse |
|  |  |  | Sally Hodgkinson | Clinical Trials Pharmacy Technician |
|  |  |  | Tanisha Patel | Pharmacist |
| Sheffield Teaching Hospitals NHS Foundation Trust | Royal Hallamshire Hospital | Sheffield | David Capener | Radiographer |
|  |  |  | Carol Carter | Pharmacist |
|  |  |  | Tracy Jackson | Pharmacy technician |
|  |  |  | Helen Bowler | Pharmacy technician |
|  |  |  | Gavin Brittain | Blinded assessor |
|  |  |  | David Paling | Co-Investigator |
|  |  |  | Basil Sharrack | Principal Investigator |
|  |  |  | Stephanie Scott | Research Nurse |
|  |  |  | Leanne Milner | Research Nurse |
|  |  |  | Sarah Johnson | Research Nurse |
|  |  |  | Carol Carter | Senior pharmacy technician |
|  |  |  | Leanne Armstrong | Radiographer |
|  |  |  | Rachel Whitham | Research Nurse |
|  |  |  | Kim Turner | Research Nurse |
|  |  |  | Katie Strobin | Pharmacy technician |
|  |  |  | Azza Ismail | Co-Investigator |
|  |  |  | Sarah Kelly | Research Nurse |
|  |  |  | Diane Sharkey | Data co-ordinator |
| University Hospitals of North Midlands NHS Trust | Royal Stoke Hospital | Stoke | Seema Kalra | Principal Investigator |
|  |  |  | Ukraina Garcia | Research practitioner |
|  |  |  | Phil Whitmore | Pharmacist |
|  |  |  | Roby Abraham | Co-Investigator |
|  |  |  | Mdtariqul Islam | Blinded assessor |
|  |  |  | Tatiana Mihalova | Blinded assessor |
|  |  |  | Susan Brammer | Pharmacist |
|  |  |  | Christine Blanks | Pharmacist |
|  |  |  | Paul Hollinshead | Clinical trials pharmacy technician |
|  |  |  | Priscilla Mhembere | Research Nurse |
|  |  |  | Mia Marsden | Clinical Research Co-ordinator |
|  |  |  | Agnes Muthoni | Research Nurse |
|  |  |  | Martin Booth | Research practitioner |
| The Newcastle upon Tyne Hospitals NHS Foundation Trust | Royal Victoria Infirmary | Newcastle | Joe Guadagno | Principal Investigator |
|  |  |  | Danielle Hall | Research Nurse |
|  |  |  | Caroline Stubbs | Research Nurse |
|  |  |  | Trish Mazambani | Research Nurse |
|  |  |  | Thomas Jarvis | Data Manager |
|  |  |  | Philip English | Radiographer |
|  |  |  | Emma Dowling | Clinical trials pharmacy technician |
|  |  |  | Julie Stephenson | Clinical trial assistant - Pharmacy |
|  |  |  | Lee Blackie | CT pharmacy support officer |
|  |  |  | Teresa Moscrop | CT pharmacy support officer |
|  |  |  | Lesley Rigden | Pharmacy technician |
|  |  |  | Hester Smith | Pharmacist |
|  |  |  | Lisa Robson | MS nurse |
|  |  |  | John Davis | Research Team Lead |
|  |  |  | Jade Deighton | Pharmacist |
|  |  |  | Martin Duddy | Blinded assessor |
|  |  |  | Hestor Rose Garratt | Blinded assessor |
|  |  |  | Vivienne Eloise Evans | Blinded assessor |
|  |  |  | Louise Thomas-Brown | MS nurse |
|  |  |  | Amal Samaraweera | Blinded assessor |
|  |  |  | Jenny Haworth | Research Nurse |
|  |  |  | Ann Hudson | Research nurse |
| University Hospital Southampton NHS Foundation Trust | Southampton General | Southampton | Prof Ian Galea | Principal Investigator |
|  |  |  | Deborah Shode | Clinical Research Fellow |
|  |  |  | Elisabeth Jarman | Research physiotherapist |
|  |  |  | Marika Sarkkinen | Research nurse |
|  |  |  | Maya Leibowitz | Blinded assessor |
|  |  |  | Mahmoud Elbahnasawi | Co-investigator |
|  |  |  | Chris Everitt | Radiographer |
|  |  |  | Ivanila Atanasova | Pharmacist |
|  |  |  | Tania Nascimento | Clinical Trials Pharmacy Technician |
|  |  |  | Elena Purcaru | MS fellow |
|  |  |  | Ratna Krishnaswamy | Clinical Research Fellow |
|  |  |  | Arshi Iqbal | Senior Research Physiotherapist |
|  |  |  | Kate Sheppard | Senior clinical trials assistant |
|  |  |  | Iberedem Imana | Senior clinical trials assistant |
|  |  |  | Veena Agarwal | Research physiotherapist |
|  |  |  | Saima Sheikh | Blinded assessor |
|  |  |  | Ivan Lumu | Clinical Research Fellow |
|  |  |  | Laura Knight | Pharmacy technician |
|  |  |  | Steve Burnage | Research Nurse |
|  |  |  | Stefania Kaninia | Clinical Research Fellow |
|  |  |  | Monica Fenn | Research Nurse |
|  |  |  | Rolando Marino | Research Nurse |
|  |  |  | Maria Bonello | Co-Investigator |
|  |  |  | Peter Fernandes | Co-Investigator |
| North Bristol NHS Trust | Southmead Hospital | Bristol | Claire Rice | Principal Investigator |
|  |  |  | Catherine Humphries | Research Nurse |
|  |  |  | Alice Ballard | Radiographer |
|  |  |  | Simon Holloway | Pharmacy technician |
|  |  |  | Sharon Hook | Pharmacy technician |
|  |  |  | Joana Cancino | Pharmacy technician |
|  |  |  | Ann Chaloner | Pharmacy technician |
|  |  |  | Lauren Pattemore | Research Nurse |
|  |  |  | Beverley Faulkner | Research Nurse |
|  |  |  | Debbie Frost | Research Nurse |
|  |  |  | Kyle James | Pharmacy technician |
|  |  |  | Eleanor Helps | Pharmacy technician |
|  |  |  | Lu Yang | Blinded assessor |
| North Bristol NHS Trust | Southmead Hospital | Bristol | Elizabeth Barnett | Research Nurse |
|  |  |  | Gayathri Subramani | Research administrator |
|  |  |  | Christopher Paisey | Blinded assessor |
|  |  |  | Deanna Stephens | Blinded assessor |
|  |  |  | Fiona Sutton | Co-Investigator |
|  |  |  | Richard Capes | Co-Investigator |
|  |  |  | Safiya Zaloum | Blinded assessor |
|  |  |  | Amy Patel | Co-investigator |
| Lothian Health Board | The Anne Rowling Regenerative Neurology Clinic | Edinburgh | Peter Foley | Principal Investigator |
|  |  |  | Dawn Lyle | Research nurse |
|  |  |  | Jill Hunter | Research nurse |
|  |  |  | Michaela Stuart | Research nurse |
|  |  |  | Judith Watt | Research nurse |
|  |  |  | Isaac Chau | Research nurse |
|  |  |  | Iona Hamilton | Radiographer |
|  |  |  | Lynsey Bagshaw | Pharmacist |
|  |  |  | Dianne Beaton | Research administrator |
|  |  |  | Don Mahad | Co-Investigator |
|  |  |  | Hatice Bozkurt | Clinical Research Fellow |
|  |  |  | Angela Crawford | Pharmacist |
|  |  |  | Ethan Stoker | Research nurse |
|  |  |  | Georgia Andreopoulou | Blinded assessor |
|  |  |  | Jessica Gill | Research Intern |
|  |  |  | Micheala Johnson | Research Intern |
|  |  |  | Sai Arathi Parepalli | Clinical Research Fellow |
|  |  |  | Riana Ali | Research practitioner |
|  |  |  | Dominic Ng | Clinical Research Fellow |
|  |  |  | Hanne Haagenrud | Senior research nurse |
|  |  |  | Lucy McLennan | Research practitioner |
|  |  |  | Chloe Parker | Research practitioner |
|  |  |  | Johnny Tam | Clinical Research Fellow |
|  |  |  | Judith Newton | Blinded assessor |
|  |  |  | Tia Cainer | Research practitioner |
|  |  |  | Daniel Sandler | Clinical Research Fellow |
|  |  |  | Rioghnach Hannan | Clinical Research Fellow |
|  |  |  | Hatice Kurucu | Clinical Research Fellow |
|  |  |  | Alexandra Entwistle-Thompson | Clinical Research Fellow |
|  |  |  | Kay (Michaela) Johnson | Clinical Research Co-ordinator |
|  |  |  | Robin Pillinger | Clinical Research Co-ordinator |
|  |  |  | Tanya van der Westhuizen | Clinical Research Co-ordinator |
| University College London Hospital NHS Foundation Trust | UCLH | London | Jeremy Chataway | Principal Investigator |
|  |  |  | Charles Wade | Co-Investigator |
|  |  |  | Sean Apap Mangion | Co-Investigator |
|  |  |  | Batoul Fneich | Research nurse |
|  |  |  | Iwona Pisarek | Study Coordinator |
|  |  |  | Ana Herrera Jimenez | Research nurse |
|  |  |  | Eirini Samdanidou | Research nurse |
|  |  |  | Jessica Roberts | Co-Investigator |
|  |  |  | Sebastian Sterkowicz | Support assistant |
|  |  |  | Marios Yiannakas | Radiographer |
|  |  |  | Anestis Passalis | Radiographer |
|  |  |  | Nathalie Passeron | Research pharmacist |
|  |  |  | Marie Braisher | Research Manager |
|  |  |  | Floriana de Angelis | Co-Investigator |
|  |  |  | Sarah Wright | Co-Investigator |
|  |  |  | Catherine Smith | Blinded assessor |
|  |  |  | Karen Gunanayagam | Blinded assessor |
|  |  |  | Sachini Kaldera | Research nurse |
|  |  |  | Luis Kirk | Pharmacy technician |
|  |  |  | Tristan Urdangarin | Pharmacist |
|  |  |  | Catarina Soares | Pharmacist |
|  |  |  | Ayse Gertude Yenicelik | Blinded assessor |
|  |  |  | James Braisher | Data Manager |
|  |  |  | Megan Wynne | Research nurse |
|  |  |  | Alessia Bianchi | Blinded assessor |
|  |  |  | Fatima Pansari | Radiographer |
|  |  |  | Neena Kim | Blinded assessor |
|  |  |  | Michelle Baird | Pharmacist |
|  |  |  | Nikita Parmar | Pharmacist |
|  |  |  | Scilla Secchi | Pharmacist |
|  |  |  | Bhavna Kaul | Blinded assessor |
| University Hospitals Coventry & Warwickshire NHS Trust | University Hospital Coventry & Warwickshire | Coventry | Tarunya Arun | Principal Investigator |
|  |  |  | Karen Jane Smallman | Research nurse |
|  |  |  | Kelly Westwood | Research nurse |
|  |  |  | Gail Evans | Research nurse |
|  |  |  | Michael Diokno | Radiographer |
|  |  |  | Mojid Khan | Pharmacist |
|  |  |  | Aminat Avosuah Seidu | Radiographer |
|  |  |  | Kirandeep Pachoo | Pharmacy technician |
|  |  |  | Stacey Clarke | Clinical trials pharmacy technician |
|  |  |  | Rajesh Varghese | Clinical trial assistant - Pharmacy |
|  |  |  | Padama Singh | Clinical trials pharmacist |
|  |  |  | Angeka Mulqueen | Clinical trial assistant - Pharmacy |
|  |  |  | Lucy Miller | Pharmacist |
|  |  |  | Abdullah Shehur | Co-Investigator |
|  |  |  | Saahitya Subramanian | Clinical Trials Pharmacy assistant |
|  |  |  | Philippa Tilby | Senior trials pharmacist |
|  |  |  | Sarmad Al-Araji | Blinded assessor |
|  |  |  | Andrew Davenport | Research charge nurse |
|  |  |  | Dashne Omar | Co-Investigator |
|  |  |  | Alexi-Jai Brown | Administrator |
|  |  |  | Sophie Weir | Pharmacy technician |
|  |  |  | Victoria Salako | Research nurse |
|  |  |  | Jaymie Johnston | Blinded assessor |
|  |  |  | Di Liang | Blinded assessor |
|  |  |  | Samir Pal | Pharmacist |
|  |  |  | Keeya Bottomley | Pharmacist |
|  |  |  | Sukhbinder Salh | Clinical trials pharmacist |
|  |  |  | Faith Omoregie | Research nurse |
|  |  |  | Luanne Carey | Pharmacy technician |
|  |  |  | Rachel Thompson | Clinical Trials Pharmacy Technician |
|  |  |  | Gaminder Shankrowala | Clinical Research Co-ordinator |
|  |  |  | Harriet Cummins | Clinical Trial assistant |
| Cardiff and Vale University Local Health Board | University Hospital Wales | Cardiff | Emma Tallantyre | Principal Investigator |
|  |  |  | Cynthia Butcher | Research Nurse |
|  |  |  | Belinda Gunning | Research Nurse |
|  |  |  | Elizabeth Perreira | Research Nurse |
|  |  |  | Kathryn Murray | Pharmacist |
|  |  |  | Luke White | Radiographer |
|  |  |  | Catherine Thomas | Radiographer |
|  |  |  | James Davies | Radiographer |
|  |  |  | Rhiannon Swanson | Clinical trials pharmacy technician |
|  |  |  | Samantha Loveless | Biorepository Manager |
|  |  |  | Andrew Thomas | Biorepository technician |
|  |  |  | Valerie Anderson | Research administrator |
|  |  |  | Jayne Howlett | Research administrator |
|  |  |  | Annie Rainey | Clinical trials pharmacy technician |
|  |  |  | Barbara Redmond | Clinical trial assistant - Pharmacy |
|  |  |  | Sian Widdows | Pharmacist |
|  |  |  | Zin Min Htet | Blinded assessor |
|  |  |  | Neil Robertson | Co-Investigator |
|  |  |  | Mark Willis | Co-Investigator |
|  |  |  | S M Saiful Bari | Blinded assessor |
|  |  |  | Fadi Ahmed | Blinded assessor |
|  |  |  | Heather Bradburn | Pharmacy technician |
|  |  |  | Megan Voisey | Research Nurse |
|  |  |  | Karim Kreft | Co-Investigator |
|  |  |  | Aung Min Saw | Blinded assessor |

**Australian sites**

| **Hospital** | **City** | **Name** | **Role** |
| --- | --- | --- | --- |
| Griffith University (Gold Coast University Hospital) | Gold Coast | Simon Broadley | Principal Investigator |
|  |  | Sabrina Oishi | Clinical Trial Manager |
|  |  | Maggie O'Hara | Research nurse |
|  |  | Danielle Lee | Pharmacist |
|  |  | Kayla Ward | Co-investigator |
|  |  | Molly Reynolds | Co-investigator |
|  |  | Antonia McLean | Blinded assessor |
|  |  | Lidia Madrid San Martin | Monitor |
| Menzies Institute of Medical Research (University of Tasmania) | Hobart | Bruce Taylor | Principal Investigator |
|  |  | Kaylene Young | Biorepository scientist |
|  |  | Julie Campbell | Health Economist |
| Brain and Mind Centre (University of Sydney) | Sydney | Michael Barnett | Principal Investigator |
| Royal Melbourne Hospital | Melbourne | Tomas Kalincik | Principal Investigator |
| John Hunter Hospital | Newcastle | Jeannette Lechner-Scott | Principal Investigator |
| The Alfred (Monash University) | Melbourne | Helmut Butzkueven | Principal Investigator |
|  |  | Vilija Jokubaitis | Biorepository scientist |
| Perron Institute | Perth | Allan Kermode | Principal Investigator |
| Concord Hospital | Sydney | Sudarshini Ramanathan | Principal Investigator |
| C/O MS Australia | Sydney | Andrew Potter | pwMS |

**MRC CTU at UCL OCTOPUS Trial team**

| **Name** | **Role** |
| --- | --- |
| Cheryl Pugh | Clinical Project Manager |
| Elizabeth Brodnicki | Clinical Trial Manager |
| Monica Lewis | Clinical Trial Manager |
| Farjana Haque | Clinical Trial Manager |
| Shabinah Ali | Clinical Trial Manager |
| Aoife Nolan | Clinical Trial Manager |
| Charlotte McGowan | Clinical Trial Manager |
| Brendan Murphy | Data Manager |
| Shuchi Naik | Data Manager |
| Sara Peres | Data Manager |
| Christos Maniatis | Data Manager |
| Olivia Mahoro | Data Manager |
| Daneil Clarke | Data Manager |
| Hannah Sweeney | Data Management Specialist |
| Maggie Hook | Data Management Specialist |
| Mary Rauchenberger | Database Programmer |
| Yinka Sowunmi | Database Programmer |

### 2. Assessment Schedule (Table 1)

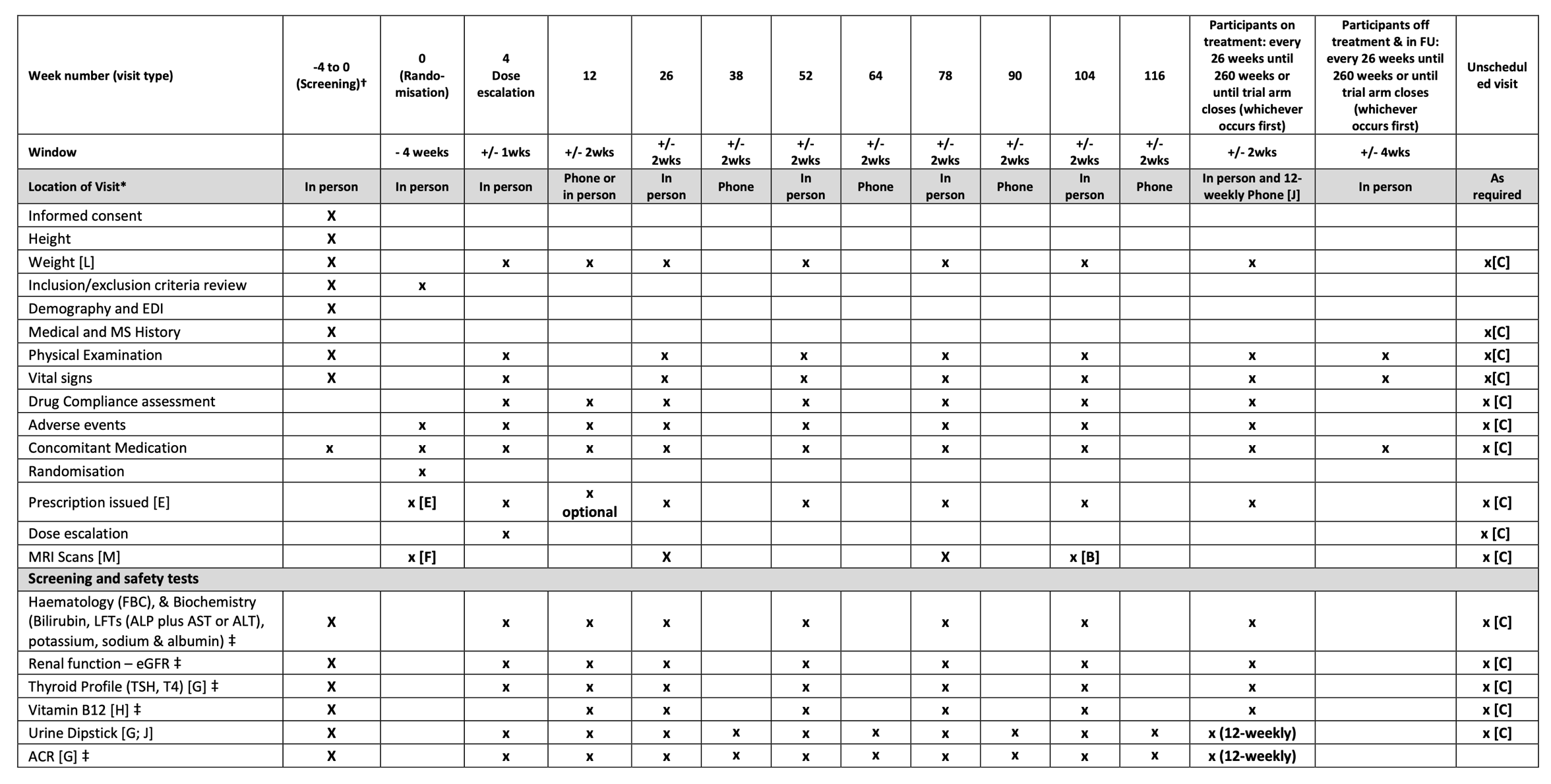

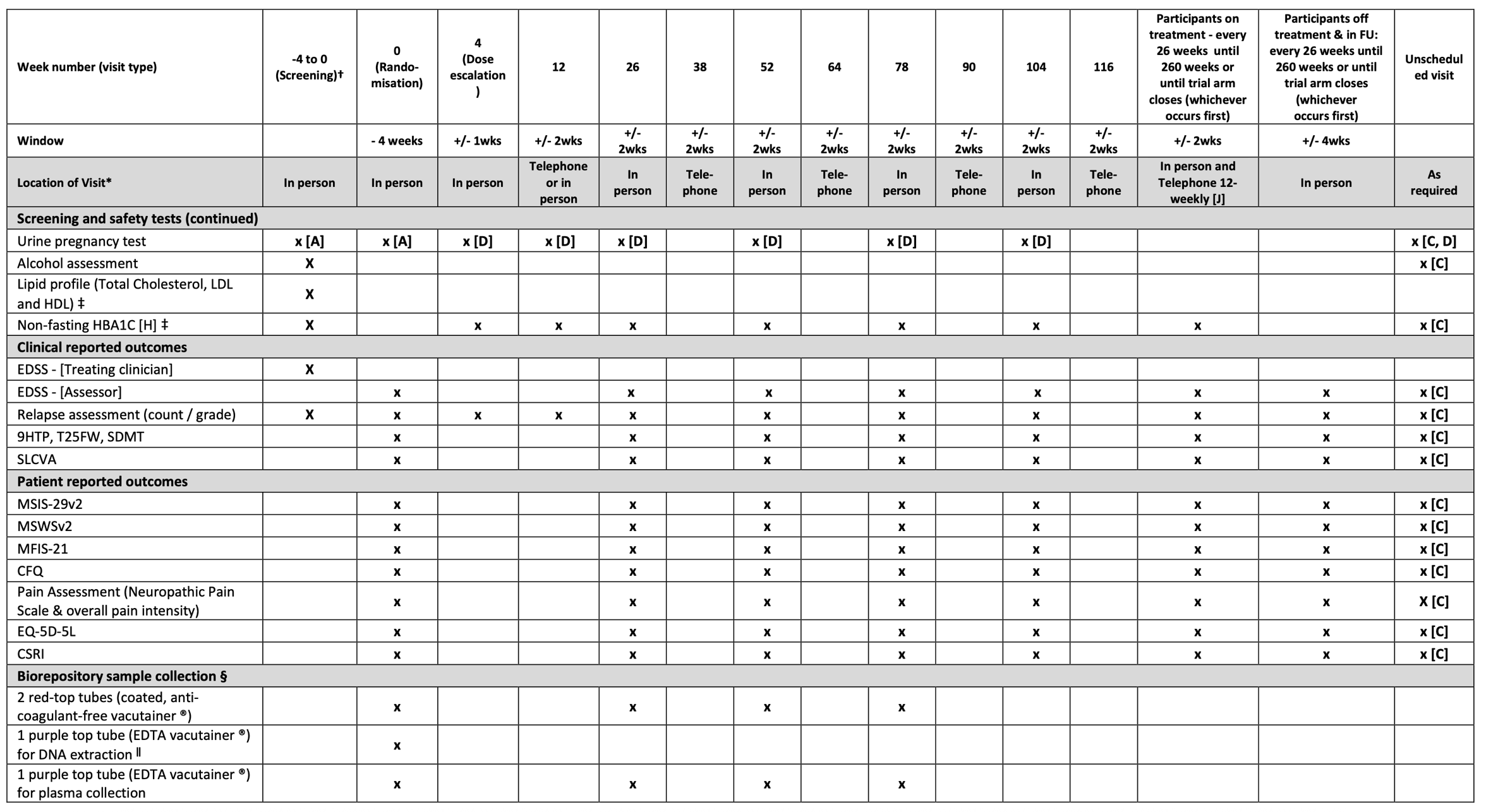

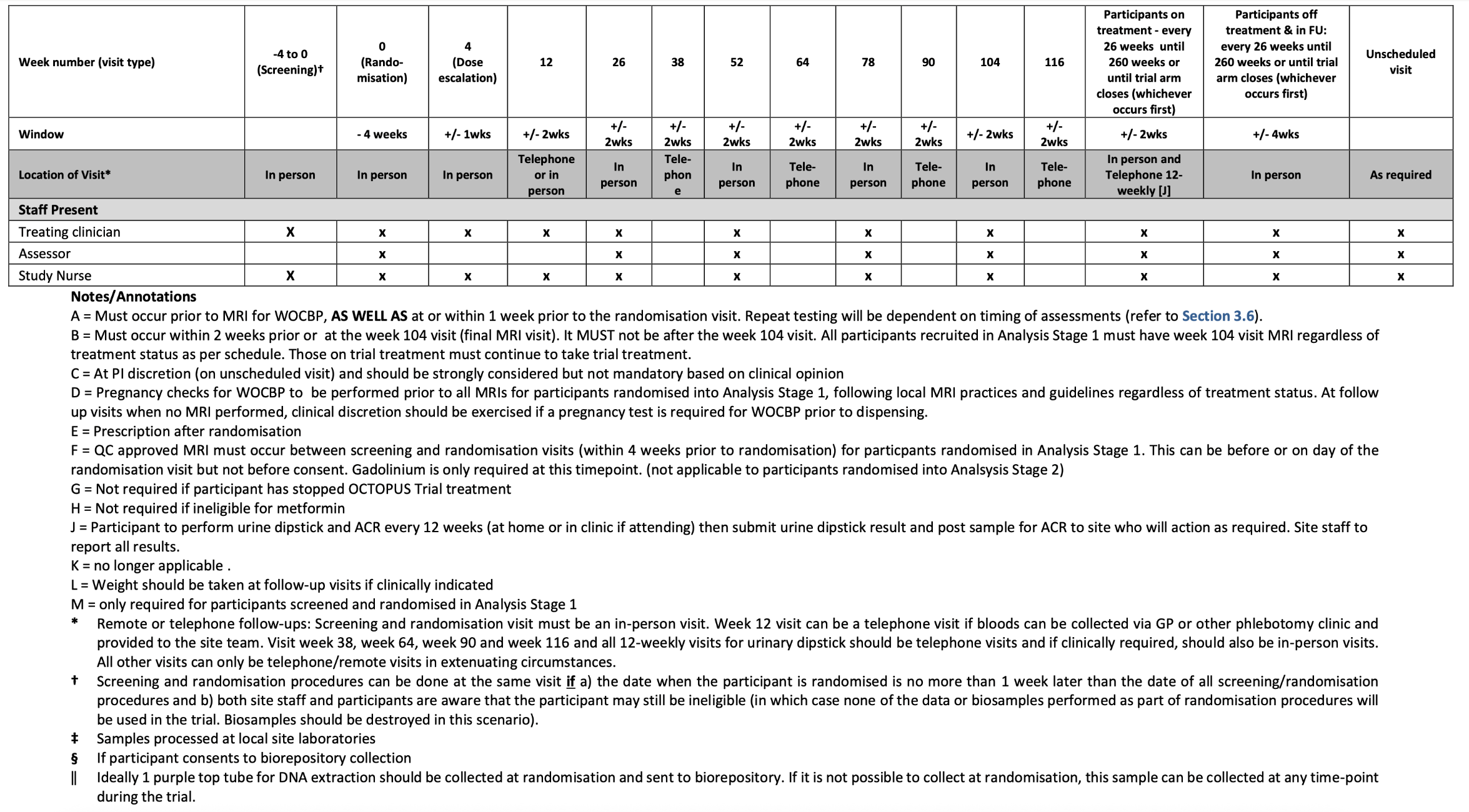

### 3. OCTOPUS Trial Committees (Figure 1)

There will be several committees involved with the oversight of OCTOPUS.

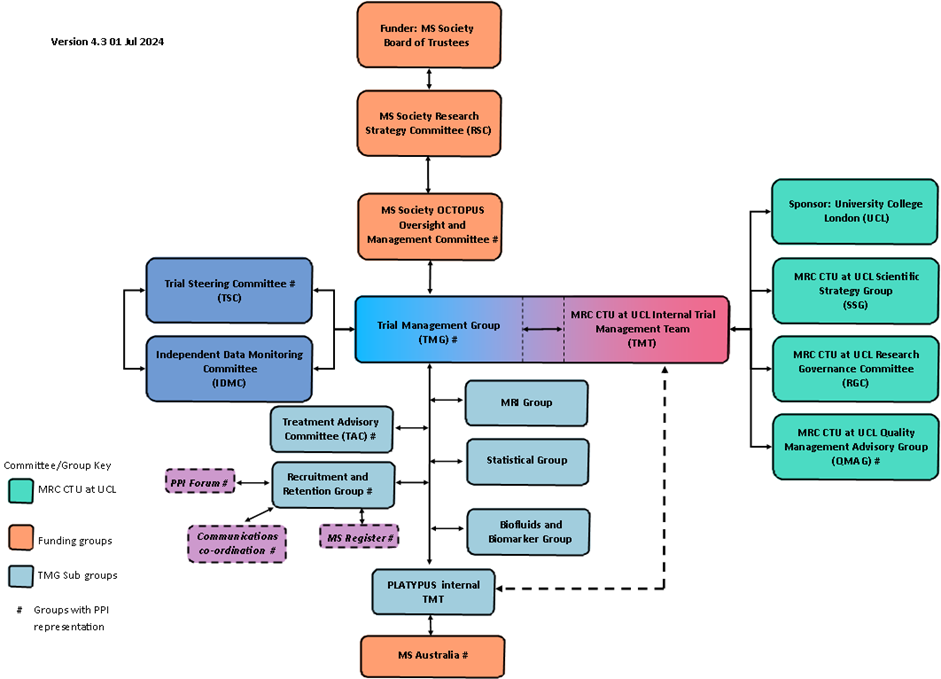

*Structure of the OCTOPUS oversight committees. MRC CTU, Medical Research Council Clinical Trials Unit; MS, Multiple sclerosis; PPI, participant and public involvement; UCL, University College London*

A Trial Steering committee (TSC) and Independent Data Monitoring committee (IDMC) will be formally responsible for the oversight of the trial, ensuring it is conducted in compliance with Good Clinical Practice (GCP), relevant regulations and ethics committee permissions. The IDMC will see the confidential, accumulating data for the trial and, using the IDMC charter and statistical analysis plan, advise the TSC. The ultimate decision for trial/arm continuation or closure will then lie with the TSC in consultation with the Trial Management Group (TMG).

The TMG will be responsible for operational oversight and management, while the Trial Management Team (TMT) will perform the day-to-day running of the trial. The TMG, comprising the Chief Investigator (CI), lead investigators (clinical and nonclinical), members of the MRC Clinical Trials Unit (CTU) OCTOPUS Team, PPIE contributors and a representative of the funder (UK MS Society), can be further divided into a number of TMG sub-specialty groups (e.g. Recruitment and Retention, and MRI). The TMT, led by the MRC CTU at UCL, will also comprise several sub-working groups (e.g. scientific strategies, research and publication). Membership (current and previous) of these groups and committees are listed below.

UCL is the Sponsor for the trial, with delegated authority to the MRC CTU at UCL.

| **Committee** | **Name** | **Role/Title** | **Organisation** |
| --- | --- | --- | --- |
| TMG | Jeremy Chataway | TMG chair/CI; Professor of Neurology | UCL QSMSC, Institute of Neurology |
| TMG | Alan Thompson | Dean of Faculty of Brain Sciences | Faculty of Brain Sciences, UCL |
| TMG | Frederik Barkhof | Professor of Neuroradiology | UCL QSMSC, Institute of Neurology |
| TMG | Siddharthan Chandran | Professor of Neurology | University of Edinburgh |
| TMG | Sue Pavitt | Professor in Translational & Applied Health Research | University of Leeds and NIHR |
| TMG | Denise Fitzgerald | Professor of Neuroimmunology | Queens’ University Belfast |
| TMG | Anna Williams | Professor of Regenerative neurology | Edinburgh University |
| TMG | Marie Braisher | Research Manager | UCL QSMSC, Institute of Neurology |
| TMG | Susan Scott | PPI advisor |  |
| TMG | Jenny Nicholas | Associate Prof of medical statistics (unblinded Statistician) | LSHTM |
| TMG | Fiona Magill | Research Nurse | Belfast City Hospital |
| TMG | Emma Gray | Assistant Director of Research | MS Society |
| TMG | Emma Tallantyre | Clinical Senior Lecturer | Cardiff University |
| TMG | Amanda Adler | Professor of Diabetic Medicine and Health Policy | Churchill Hospital, Oxford |
| TMG | Max Parmar | Vice Chair / Director | MRC CTU at UCL |
| TMG | Matthew Burnell | Trial Statistician | MRC CTU at UCL |
| TMG | Cheryl Pugh | Clinical Project Manager | MRC CTU at UCL |
| TMG | Monica Lewis | Clinical Trial Manager | MRC CTU at UCL |
| TMG | Liz Brodnicki | Clinical Trial Manager | MRC CTU at UCL |
| TMG | Farjana Haque | Clinical Trial Manager | MRC CTU at UCL |
| TMG | Sam Loveless | OCTOPUS Biorepository Manager | Cardiff University |
| TMG | Brendan Murphy | Data Manager | MRC CTU at UCL |
| TMG | Shuchi Naik | Data Manager | MRC CTU at UCL |
| TMG | Daneil Clarke | Data Manager | MRC CTU at UCL |
| TMG | Simon Broadley | Australia Lead Investigator | Griffith University |
| TMG | Rachel Burton | Trial Statistician | MRC CTU at UCL |
| TMG | Rod Middleton | MS Register Project Manager | Swansea University |
| TMG | James Carpenter | Senior Statistician | MRC CTU at UCL |
| TMG | Anna Combes | Research Fellow | UCL QSMSC, Institute of Neurology |
| TMG | Judith Grubb | PPI representative | N/A |
| TMG | Rachael Hunter | Health Economist (UK) | UCL |
| TMG | Ekaterina Bordea | Health Economist (UK) | UCL |
| TMG | Sean Apap Mangion | Clinical Fellow | UCL QSMSC |
| TMG | Charlie Wade | Clinical Fellow | UCL QSMSC |
| TMG | Shabinah Ali | Clinical Trial Manager | MRC CTU at UCL |
| TMG | Fleur Hudson | Head of TSM at MRC CTU | MRC CTU at UCL |
| TMG | Charlotte McGowan | Clinical Trial Manager | MRC CTU at UCL |
| TMG | Sara Peres | Data Manager | MRC CTU at UCL |
| TMG | Christos Maniatis | Data Manager | MRC CTU at UCL |
| TMG | Jacqui-Ann Hanley | Head of Research/Observer | MS Society |
| TMG | Olivia Mahoro | Data Manager | MRC CTU at UCL |
| TMG | Aoife Nolan | Clinical Trial Manager | MRC CTU at UCL |
| TMG | Michelle Naughton | Research Fellow | Queens’ University Belfast |
| TMG | Julie Campbell | Health Economist (Australia) | Menzies Institute for Medical Research, University of Tasmania |
| TMG | Deborah Shode | Clinical Fellow - PI Associate Scheme | Southampton General Hospital |
| TMG | Clare Walton | Head of Research/Observer | MS Society |
| TMG | Dawn Lyle | Research Nurse | Anne Rowling Regenerative Neurology Clinic |
| IDMC | Tim Maughan | Chair | Oxford Institute for Radiation Oncology |
| IDMC | Mia Pia Sormani | IDMC member | University of Genoa |
| IDMC | Andrew Goodman | IDMC member | University of Rochester Medical Center |
| IDMC | Chris McGuigan | Statistician | St Vincent's University Hospital, Dublin |
| IDMC | Matthew Burnell | Unblinded Statistician | MRC CTU at UCL |
| IDMC | Jenny Nicholas | Unblinded Statistician | LSHTM |
| IDMC | Mahesh Parmar | Blinded statistician | MRC CTU at UCL |
| IDMC | Rachel Burton | Unblinded Statistician | MRC CTU at UCL |
| TSC | Andrew Gale | Neurologist | Not required |
| TSC | Catherine Hewitt | Chair | Not required |
| TSC | Judith Mathie | PPI representative | Not required |
| TSC | Kate Petheram | Neurologist | Not required |
| TSC | Sarah Weller | PPI representative | Not required |
| Communications | Caitlin Astbury | Chair/Research Communications Manager | MS Society |
| Communications | Susan Scott | PPI advisor | N/A |
| Communications | Emma Tallantyre | Clinical Senior Lecturer | Cardiff University |
| Communications | Pat Poole | PPI advisor | N/A |
| Communications | Annabelle South | Policy, Communications & Research Impact Coordinator | MRC CTU at UCL |
| Communications | Eleanor Hubble | Stakeholder Engagement & Data Acquisition Officer | MS Register team at Swansea University |
| Communications | Laura Belscott | PPI lead at MS Society | MS Society |
| Communications | Jan Carver | PPI advisor | N/A |
| Communications | Berta Terre Torras | Science Communications Officer | MRC CTU at UCL |
| Communications | Catherine Godbold | Research Communications Manager | MS Society |
| Communications | Anneesa Amjad | PPI lead at MS Society | MS Society |
| Communications | Claire Walton | Head of Research/Observer | MS Society |
| Communications | Helen Burchmore | PPI advisor | N/A |
| Communications | Katie Tuite-Dalton | Stakeholder Engagement & Data Acquisition Officer | MS Register team at Swansea University |
| Communications | Becca Lawther | PPI advisor | N/A |
| Communications | Charlotte Hartley | Science Communications Officer | MRC CTU at UCL |
| MS Register | Emma Tallantyre | Chair/Clinical Senior Lecturer | Cardiff University |
| MS Register | Marie Braisher | Research Manager | UCL QSMSC, Institute of Neurology |
| MS Register | Rod Middleton | Principal Investigator and System Architect of the UK MS Register | MS Register team at Swansea University |
| MS Register | Susan Scott | PPI advisor | N/A |
| MS Register | Becca Lawther | PPI advisor | N/A |
| MS Register | Sean Apap Mangion | Clinical Fellow | UCL QSMSC |
| MS Register | Charlie Wade | Clinical Fellow | UCL QSMSC |
| MS Register | Eleanor Hubble | Stakeholder Engagement & Data Acquisition Officer | MS Register team at Swansea University |
| MS Register | Leah Jones | Stakeholder Engagement & Data Acquisition Officer | MS Register team at Swansea University |
| MS Register | Elaine Craig | Research Analyst | MS Register team at Swansea University |
| MS Register | Katie Tuite-Dalton | Stakeholder Engagement & Data Acquisition Officer | MS Register team at Swansea University |
| MS Register | Chris Brownlee | PPI advisor | N/A |
| Recruitment and retention | Jeremy Chataway | CI | UCL QSMSC, Institute of Neurology |
| Recruitment and retention | Emma Gray | Assistant Director of Research | MS Society |
| Recruitment and retention | Emma Tallantyre | Clinical Senior Lecturer | Cardiff University |
| Recruitment and retention | Cheryl Pugh | Clinical Project Manager | MRC CTU at UCL |

Please note members of the MRC CTU at UCL are also members each of the above groups/committees as required

### 4. Protocol Updates

Please note version 1.0; dated 03-Aug-2022 was not approved for use and was amended following comments from the Research Ethics Committee, MHRA and HRA. Therefore version 2.0; 11-Oct-2022 was the first approved protocol version.

**Summary of changes between v2.0 and V3.0**

In addition to correction of typos and grammatical errors, the following was updated:

- Updated MRC CTU at UCL contacts and TMG members
- Summary table - Addition of analysis stage 1 exploratory analysis
- Amendments to assessment table
  - Addition of haematology (FBC), Biochemistry (Bilirubin, LFTs (ALP plus AST or ALT)), potassium and sodium to screening and safety tests
  - Clarification that [C] is at PI discretion and should be strongly considered but not mandatory based on clinical opinion.
  - Amendment that pregnancy checks (removing pregnancy test) for WOCBP must be performed prior to all MRIs following local MRI practices and guidelines.
  - Alcohol assessments now carried out at baseline and as per clinical discretion throughout trial.
- Abbreviation additions of DSMS (Drug Supply Management System), PRLs (Paramagnetic Rim Lesions), SWI (Susceptibility weighted images) and TLC (three letter code)
- Lay summary - Updated Figure 3 inserted
- Section 1.9 - addition Exploratory Analysis: Paramagnetic Rim Lesions
- Section 3.2 - Removal of Participant Exclusion Criteria 3: Rare hereditary problems of galactose intolerance or glucose galactose malabsorption as moved to metformin specific criteria (in Metformin drug appendix)
- Section 3.6 - Addition of sentences to Section 3.6 Screening Procedures & Pre-randomisation investigations for PIS location on website, how obtain participant identification number and TC and clarifying all GPs and neurologists should be informed of participation involvement.
- Section 4 Randomisation **-** Removal of sentence for manual randomisation
- In section 5.2 Products, the wording has been clarified including confirmation that IMPs to be kept out of direct sunlight and the use of the Drug Supply Management System (DSMS)
- In section 5.5 Dispensing and Storage: clarification of use of DSMS
- In section 5.6 – confirmed *maximum dose for the remainder of the trial* will be achieved at 24 weeks and removal of sentence “no further escalations can be performed”.
- In section 5.6.1 Renal impairment – correction that all participants should pause for 24 hours prior to iodinated contrast agents and correction restart trial treatment only after eGFR has be confirmed >50ml min/1.73m^2^ (not 30ml min/1.73m^2^).
- In section 5.6.2 Gastrointestinal table 4: clarification of wording for dose modifications for gastrointestinal toxicity
- In section 5.6.3 Table 5 Management of trial treatment for proteinuria - updated to include both units for ACR.
- In section 5.6.4 Table 6 Vitamin B12 Deficiency - Addition of >200pg/ml as units and addition of sentence: If ≤200ng/l (200 pg/ml) (148pmol/l) after 9 months post initial test, discontinue trial treatment.
- In section 5.7 – clarification on pregnancy testing and checks prior to MRI scans.
- In section 5.8 Accountability and unused drugs/devices addition and clarification of wording use of the treatment log and diary cards
- In section 5.9 Compliance and Adherence clarification and rewording of the section including addition of wording on use of the electronic diary card
- Section 5.10 Handling cases of trial treatment overdose – clarification of wording to confirm if accidental overdose, participants can restart treatment, whereas for deliberate overdose treatment should stop.
- Section 5.14.3 Medications to be used with caution addition of ‘investigator brochure’ and clarification of requirements to pause or stop medications of caution.
- Section 6.2.4 Concomitant Medication addition of wording: At each visit, a review of concomitant medication must be performed to ensure any contraindicated medications including taking any Analysis Stage 1 IMPs are not being taken.
- Section 6.3 Safety Assessments addition of wording to provide specific instructions performing for the urinary dipstick and addition of the applicable colour chart.
- Section 6.3.3 Pregnancy - clarification on pregnancy testing and checks prior to MRI scans.
- Section 6.6 Procedures for assessing patient reported outcomes – clarification of wording for collection of patient reported outcomes for telephone visits and removal of the wording “The following assessments should be facilitated by the assessing clinician and/or an appropriate trial team member.”
- Sections 6.6.3 Modified Fatigue Impact Scale -21 (MFIS-21), Section 6.6.4 Chalder Fatigue Questionnaire (CFQ); Section 6.6.5 Pain Assessment and Section 6.6.6 EQ-5D-5L - removal of the wording “This should be completed at each in person follow up visit and or online via participate if participant happy to do so. This can be performed on a telephone follow up using the worksheet in extenuating circumstances”.
  - Section 9.1 Method of Randomisation clarification of the generation of the bottle numbers and to who and where they are supplied.
  - Section 16 Publication and Dissemination of Results – revision of the wording for when results of interim analyses will be available and scenarios requiring consideration.
  - Addition of metformin specific exclusion criteria: rare hereditary problems of galactose intolerance or glucose-galactose malabsorption
  - Clarification of units in R/S-Alpha lipoic acid specific exclusion eligibility criteria – Urinary dipstick for proteinuria 1+ or higher and albumin/creatinine ratio (ACR) >300mg/g or ≥ equal to 34mg/mmol

**Summary of changes between v3.0 and V4.0**

- Update to Trial team – Charlotte McGowan replaced as Trial Manager by Elizabeth Brodnicki
- Section 3.2 - Reinsertion of Participant Exclusion Criteria 3: Rare hereditary problems of galactose intolerance or glucose galactose malabsorption as previously moved to metformin specific criteria (in Metformin drug appendix)
- Update to R/S-Alpha lipoic acid specific appendix in section 2.2 the exclusion eligibility criteria clarification of units 34mg/mol should read 34mg/mmol. Therefore appendix version updated to v4.0; 07-Sept-2023. Please note the Metformin specific appendix was not updated in this amendment and remains at v3.0.

**Summary of changes between v4.0 and V5.0**

In addition to correction of typos and grammatical errors, the protocol was updated to include the participation of Australian sites, and to ensure that any UK-specific references were amended accordingly. References to local Country-Specific Appendices were added where appropriate for additional information about trial conduct specifically in the country.

- Addition of Australian logos
- Inclusion of PLATYPUS (Australian extension of the OCTOPUS trial)
- Updates to compliance section to include sites outside the UK and EU/EEA
- Addition of funding bodies for PLATYPUS, the Australian extension of the OCTOPUS trial
- Updates to safety reporting contact details to make applicable for sites outside the UK
- Trial Administration and Co-ordinating Centre sections updated to include Olivia Mahoro, Aoife Nolan, and the Australian team contact details
- Simon Broadley added to the TMG member list
- MS Australia and MSWA added as funders
- Trial schema (figure 1 in protocol and drug appendices) updated to say “National Registers” instead of “MS Register”
- The trial assessment schedule was amended as follows:
  - TSH and T4 testing specified under thyroid profile tests
  - Vitamin B12 test is no longer required at week 4 visit
  - EDSS assessing clinician is now termed “EDSS assessor”
- Lay summary updated to include references to the trial activity outside the UK (i.e. PLATYPUS activity in Australia)
- GP abbreviation updated to include “(or known as family doctor outside UK)”
- PLATYPUS, Country Coordinating centre, country specific appendix and country lead sites added to abbreviation list
- Section 2.2 Approval and Activation, and section 2.3 Site Management: references to the CSA added here for non-UK sites
- Section 3.1 Participant Core Inclusion Criteria: eGFR cut off changed from eGFR ≥60ml/min/1.73m^2^ to eGFR ≥65ml/min/1.73m^2^
- Section 3.2 Participant Core Exclusion Criteria:
  - History of alcohol or drug use limited to within the last 5 years
  - Participation in another clinical trial of IMP of medical device ≤ 26 weeks before randomisation updated to “Use of an investigational medical product or investigational medical device ≤ 26 weeks before randomisation”
- Section 3.6:
  - Reference made to screening procedures used in Australia
  - Wording updated to state that a combination of on-site and remote monitoring of the completed consent forms will be utilised through the course of the trial
- Section 5.6 Expected Toxicities, Dose Modification & Discontinuations: the following sentence was removed “Doses can then only be reduced, paused or stopped due to safety reasons, clinician choice or participant choice. This dose change must be performed by the treating clinician.”
- Section 5.6.1 Renal Impairment: parameters for renal impairment management were changed so that participants with eGFR of 45 – 59 ml/min/1.73m^2^ must re-test within 4 weeks but can continue on current dose. If result remains 45 – 59 ml/min/1.73m^2^ participants can remain on current dose but must be re-tested again at next in-person visit. Those with an eGFR <45 must permanently stop trial treatment.
- Section 5.6.3: management of proteinuria changed so that If ≥ 1000 mg/g (>113 mg/mmol), treatment must be paused until mandatory retest is completed (within 4 weeks). If retest remains ≥ 1000 mg/g (>113 mg/mmol), treatment must be stopped permanently.
- Section 5.6.4 Vitamin B12 deficiency management includes a statement confirming that it is up to clinical discretion whether to return to high dose immediately after successful B12 replacement therapy or to gradually escalate trial IMP over 2 weeks. It also highlights that a B12 test is not required at the week 4 visit
- Section 5.6.5 updated to allow for a temporary pause (not just a dose reduction) if a participant cannot tolerate high dose trial treatment
- Treatment and dose diagrams for low and high dose updated (Figure 7 and 8)
- Section 5.10 and both drug appendices: removal of the following sentence “Any dose in excess of that specified according to the protocol will constitute an overdose”.
- Section 5.11.1 Emergency Unblinding section updated to make wording more generic by removing UK/NHS-specific references. Also updated to state that it can be carried out only in a medical emergency or situation
- Section 5.14.2 Medications not permitted: Definition of excessive alcohol is now as per investigator discretion.
- Section 5.14.3 Medications to be used with caution: instructions for re-starting after pause have been updated so that trial treatment can be resumed even if they are not within 24 weeks of randomisation.
- Section 6.1: more clarity on who can carry out assessments
- Section 6.2: blood tests can now be completed up to 2 weeks prior to the treatment visit (previously this was 1 week). Procedures must be in place for cases where a dose modification or treatment change is needed following blood test review.
- Specific UK provider of the urine dipstick kits removed
- Section 6.5: EDSS assessment instructions updated to include a reference to section 6.1.
- Section 6.5.6: Wording updated to clarify that the severity of grade 1 and 2 relapses should be documented in the medical notes and added to the AE log on the database. Grade 3 relapses should be reported as an SAE
- References to the Participate module of the database added to sections 5.9, 6.6, 6.6.7 and 8.4.
- Section 7.1 Safety reporting definitions updated to refer to the Medicines for Human Use (Clinical Trials) Regulations 2004 (SI 2004/1031) and subsequent amendments, ICH E2A “Clinical Safety Data Management: Definitions and Standards for Expedited Reporting” and ICH GCP E6.
- Section 7.1.3 Adverse and disease related events exempt from expedited reporting: “medical or surgical procedures; the condition that leads to the procedure is the adverse event” was moved from the bullet point list to the body of the paragraph test above
- Section 7.3.7 Notification procedure: instructions for reporting positive pregnancy test added (enter result on Lab eCRF and not on AE log).
- Minimum criteria for reporting an SAE was amended to remove date of birth
- Section 7.4 Sponsor responsibilities (MRC CTU at UCL): process for reporting outside the UK (via a country coordinating centre) added
- Section 8.4 Source Data: instances where eCRF is source data have been added
- Section 10.1 Biorepository: Reference to the CSA included for non-UK countries.
- Section 11.1 Compliance: updated wording to reference regulatory framework introduced since last amendment. Compliance wording for non-UK sites aso added.
- Section 11.2.1 Ethical considerations: Travel expenses for participants changed from £40 to “maximum amount per visit is defined in site agreement”
- Section 11.2.2 Favourable ethical opinion: wording updated so it is also applicable to sites outside UK
- Section 11.3 Competent authority approvals: wording updated so it is also applicable to sites outside UK
- Section 11.5: Trial closure: wording updated so it is also applicable to sites outside UK
- Section 13 Finance: wording updated so it is also applicable to sites outside UK
- Section 14.1: Trial Management Group (TMG): wording updated to add reference to input from country coordinating centres.
- Figure 11 Relationship of trial committees scheme updated to include country coordinating centre input
- Section 15: Patient and Public Involvement: reference to Australia’s PPI added
- Addition of drug manufacturers to metformin and R/S-ALA appendices
- Metformin drug appendix section 3.1: Changed “1000mg tablets” to “500mg tablets”
- R/S-ALA drug appendix section 3.1: Added clarification that 300mg capsules of R/S-ALA will be over encapsulated.

**Summary of changes between v5.0 and V6.0**

- Section 3.1 Participant Core Inclusion Criteria: eGFR cut off changed from eGFR ≥65ml/min/1.73m^2^ to eGFR ≥60ml/min/1.73m^2^

**Summary of changes between v6.0 and V7.0**

The main change in this protocol amendment is the removal of the QA approved MRI inclusion criteria, and the requirement for 3 follow up MRIs for those recruited in Analysis Stage 2. Some future tenses were also amended where it refers to aspects of the trial that have already been completed. A summary of all changes is provided below:

- Trial Administration: removal of data manager Christos Maniatis and replacing with Daneil Clarke
- Trial Administration: removal of trial manager Chloe Osbourne and replacing with Vanessa Vigar
- Trial Assessment Schedule: Week number (visit type) column headings updated from “On treatment” to ”Participants on treatment”, and “Completed treatment” to “Participants treatment & in FU”
- Trial Assessment Schedule: location of visit headings changed from “Telephone” to “Phone”
- Trial Assessment Schedule: footnotes B, D, F, and M updated to highlight that MRI is only required for participants randomised into Analysis Stage 1
- Lay Summary Background: specific reference to first Analysis Stage added to the “How will this trial be carried out” section ensuring that it is clear MRIs are only applicable to this Stage
- Abbreviations: duplicate 9HPT removed
- Section 1.6.1: reference added that use of MRI is only for Analysis Stage 1
- Section 2.1.1: GCP training requirement changed from 2 years to “2 to 3 years”
- Section 2.2: Approval and Activation - reference added to state that an MRI QA monitoring system is required for sites participating in Analysis Stage 1 only
- Section 2.3 Site Management – reference to Analysis Stage 1 added for QSMSC Institute of Neurology MRI responsibilities
- Section 3: wording updated to state that the eligibility criteria are for Analysis Stage 2 only (previously Analysis Stage 1)
- Section 3.1: Participant Core Inclusion Criteria – removal of criteria 10 and 11, and addition of a note highlighting that these are no longer core inclusion criteria in Analysis Stage 2:
  - [Please note no longer core inclusion criteria in Analysis Stage 2 - Must have a QC‑approved (as defined in MRI guide) MRI ≤ 4 weeks before randomisation]
  - [Please note no longer core inclusion criteria in Analysis Stage 2 - Willing and able to have MRI scans in accordance with the assessment schedule and no contraindication to MRI (please refer to MRI Procedures and Protocol for further detail)]
- Section 3.2: Participant Core Exclusion Criteria – updated wording of criteria 14 to refer to IMPs as “OCTOPUS” IMPs rather than “Analysis Stage 1” IMPs
- Section 3.6: Screening Procedures and Pre-Randomisation Investigations – updated wording to make completion of registration of interest mandatory regardless of how potential participants are identified
- Section 4.3: Re-Randomisation into OCTOPUS – added the wording “after the decision point”, and added confirmation that participants who have withdrawn cannot be re-randomised if their arm hasn’t closed
- Section 5.6.3: Proteinuria – Table 5 updated to remove “on repeat urinary dipstick”
- Section 5.6.3: Proteinuria – Table 5 footnote added to highlight that participants can only be re-challenged a maximum of two times
- Section 5.6.5: Other Toxicities – updated two typos (“capsule” to “capsules”)
- Section 5.7: Contraception – added that only WOCBP screened and randomised in Analysis Stage 1 will require pregnancy checks prior to MRI
- Section 5.9: Compliance & Adherence – corrected typo (“a link will be sent a link” to “Participants will be sent a link”) and changed timeline from “30 days” to “29 days” post all other clinic visits to complete
- Section 5.11.1: Emergency Unblinding – added sentence stating that full details and guidance for unblinding are available on the OCTOPUS website
- Section 5.13: Treatment Data Collection – added reference to Analysis Stage 1
- Section 5.14.2: Not permitted medications – changed “Analysis Stage 1 IMPs” to “OCTOPUS IMPs”
- Section 6.1: Trial Assessment Schedule – added space in third paragraph (“assessors should”)
- Section 6.3.1: Bloods – Added sentence confirming that once participants have stopped trial treatment, bloods are not mandatory for the trial
- Section 6.3.3: Pregnancy – added specific reference that participants recruited in Analysis Stage 1 will require pregnancy check prior to MRIs, and clarification that pregnancies in all Analysis Stages will be reportable and must stop trial treatment
- Section 7.3.7: Notification Procedure – “The SAE or NE must entered to…” amended to “The SAE or NE must be entered onto the…”
- Section 10.2: Biorepository Governance – updated to include reference to Australian biosamples and that governance and ownership of these samples will be transferred to Griffith University (Australian National Sponsor) after the end of the trial. It was also noted that proposals for future use of Australian biosamples will be reviewed by Griffith University on behalf of the OCTOPUS TMG
- 11.2.1: Ethical Considerations – reference to Analysis Stage 1 added to second bullet point
- Section 13: Finance – reference to Analysis Stage 1 added for MRI scans acquisition costs
- Section 14.1: TMG – minor update to MRI Group description
- Section 15.3: Identifying PPI Contributors – updated wording to state that the team aims to include a minimum of five people affected by MS as members of the TMG (including subgroups), and removed reference to the MRI group in this section

**Summary of changes between v7.0 and V8.0**

The main change in this protocol amendment is the addition of mandatory albumin-creatinine-ratio (ACR) testing at screening and throughout the trial. This includes implementation of at-home urine sample collections for interim testing between in-person visits. A number of additional exclusion criteria have also been added to ensure the safety of potential trial participants. A summary of all changes is provided below:

- Trial Administration: addition of statistician Rachel Burton
- Trial Administration: removal of data manager Olivia Mahoro and replacing with Brendan Murphy
- Trial Administration: removal of trial manager Vanessa Vigar and replacing with Sabrina Oishi
- Trial Assessment Schedule: week number (visit type) column headings updated for consistency across pages
- Trial Assessment Schedule: prescription at week 12 updated to “optional”
- Trial Assessment Schedule: albumin added to haematology and biochemistry test list
- Trial Assessment Schedule: biorepository sample collection timepoints updated to remove collections beyond week 78
- Trial Assessment Schedule: footnotes B, F, G, J and K updated to clarify requirements around week 104 MRI and urine testing
- Section 3: Selection of Participants - clarification added to state that eligibility criteria only apply at the point of randomisation
- Section 3.2: Participant Core Exclusion Criteria –
  - addition of “significant non-MS neurological comorbidity” to criterion 2
  - updated wording of criterion 12 to refer to IMPs as “OCTOPUS” IMPs rather than “Analysis Stage 1” IMPs
  - updated wording of criterion 13 to include 3,4-aminopyridine
  - addition of criterion 16 to exclude participants that have had previous treatment with alemtuzumab or autologous haematopoietic stem cell therapy (AHSCT) ≤ 52 weeks prior to randomisation
  - addition of criterion 17 to exclude participants with an Albumin Creatinine Ratio (ACR) result of ≥34 mg/mmol (>300 mg/g) at screening, regardless of urine dipstick results
  - addition of criterion 18 to exclude participants with a diagnosis of diabetes mellitus
- Section 3.5: Co-Enrolment Guidelines – updated reference to section 4.2
- Section 3.6: Screening Procedures and Pre-Randomisation Investigations – changed “PLATYPUS Consortium Portal” to “MS Trial Screen”
- Section 3.6: Screening Procedures and Pre-Randomisation Investigations – updated wording to include reference to urine testing
- Section 3.6: Screening Procedures and Pre-Randomisation Investigations – addition of reference to the Australian Country Specific Appendix for local Australian requirements
- Section 5.4.1: Initial or Low Dose – participants must take two capsules once daily for a minimum of 3 weeks following randomisation before consideration for dose escalation
- Section 5.4.2: High Dose – updated so that the first 26 weeks (rather than 24 weeks) post randomisation will determine the participants maximum dose during the trial
- Section 5.6: Expected Toxicities, Dose Modifications & Discontinuations – updated from 24 to 26 weeks
- Section 5.6.3: Proteinuria – section updated to reflect new requirement for ACRs to be carried out every 12 weeks, alongside urine dipsticks
- Section 5.6.3: Proteinuria – Table 5 updated to reflect new ACR requirements. It also includes guidance for trial teams to refer participant to local nephrology team for evaluation if ACR result is above normal range
- Section 5.6.3: Proteinuria – Table 5 footnote added to highlight that all cases of macroalbuminuria, nephrotic syndrome, and glomerulonephritis should also be reported as notable events (NEs) and that any renal biopsy results must be noted on the related NE and AE report
- Section 5.6.4: Vitamin B12 Deficiency – Table 6 updated to refer to local lab lower limit of normal range rather than an absolute protocol cut-off range
- Section 5.6.5: Other Toxicities – guidance has been updated around managing toxicities arising when on low dose
- Section 5.6.5: Other Toxicities – Figure 7 “Treatment and dose diagram – low dose” updated to reflect updated guidance
- Section 5.12: Trial Treatment Discontinuation – addition of “commencement of any diabetic medication”
- Section 5.14.2: Medications Not Permitted – Table 7 and guidance text updated to note that diabetes medication (including insulin and metformin) are not permitted during the trial. If a participant needs to commence a non-permitted medication then they must discontinue trial treatment for the remaining duration of the trial but should remain in trial follow-up and complete all clinical assessments
- Section 5.14.3: Medications to be Used with Caution – Table 8 updated to remove diabetic drugs
- Section 6.3.2: Urine Dipstick and ACR – section updated to include at-home testing instructions for additional ACR tests required alongside urine dipstick testing
- Section 6.3.2: Urine Dipstick and ACR – Figure 10 updated to reflect new at-home ACR testing requirements
- Section 7.2.1: Toxicities – “glomerulonephritis” notable event has been expanded to define macroalbuminuria and nephrotic syndrome reporting requirements
- Section 7.2.2: Pregnancy – removal of “end of treatment period or end of trial regardless of the outcome” as pregnancies occurring during the trial will always be followed until the outcome of pregnancy has been established
- Section 10.1: Biorepository – updated to note that samples will only be collected up to and including week 78
- 11.1.3: Data Collection & Retention (Archiving) – addition of Sponsor retention requirements

Section 14: Oversight & Trial Committees - addition of reference to the Australian Country Specific Appendix for local Australian oversight committees

- Section 15.2: Patient and Public Involvement Advisory Groups – updated wording describes the PPI representatives that are currently part of the TMG, communications sub-group, and MS register sub-group
